# Molecular mechanisms associated with multiple sclerosis progression, severity and phenotype

**DOI:** 10.1101/2022.10.14.22281095

**Authors:** Peter Kosa, Keith Lumbard, Jing Wang, C. Jason Liang, Ruturaj Masvekar, Yujin Kim, Mihael Varosanec, Lori Jennings, Bibiana Bielekova

## Abstract

While current treatments of multiple sclerosis (MS) effectively inhibit formation of focal lesions and relapses, most patients experience progression independent of relapse activity (PIRA). To understand PIRA, we analyzed nine prospectively acquired clinical and imaging outcomes in 176 relapsing-remitting and 215 progressive MS patients and 45 healthy volunteers, along with matched cellular and >5000 protein data in 1,042 cerebrospinal fluid (CSF) samples. Regressing out physiological aging and sex effects identified MS-related processes. Among these, compartmentalized inflammation and its effector mechanisms such as pyroptosis showed the strongest association with MS severity, irrespective of clinical categorization of patients. However, molecular processes affected localization of CNS injury: patients with predominant brain damage had proportionally higher neuroinflammation, while fibrosis and tissue hypoxia were linked to principal involvement of spinal cord. We did not identify inflammation-unrelated neurodegeneration; instead, CNS-related processes were beneficial, such as synaptogenesis. Machine learning-based CSF biomarker models predicted nine clinical and volumetric imaging outcomes in the independent cohort with accuracy exceeding published MS models.

These data show intra-individual diversity of putative disease mechanisms in MS and implicate processes related to compartmentalized neuroinflammation as leading candidate mechanisms of PIRA. Future drug development should include CNS-penetrant anti-inflammatory agents.

## INTRODUCTION

It has been difficult to study pathogenic mechanisms of the central nervous system (CNS) diseases in living humans, because they evolve behind the blood-brain barrier (BBB). Instead, understanding of neurological diseases mostly comes from postmortem human pathology studies complemented by animal models.

This is exemplified by multiple sclerosis (MS), a polygenic inflammatory and demyelinating disease of the CNS. Perivascular inflammation in acute MS lesions, combined with studies of experimental autoimmune encephalomyelitis (EAE), recognized migration of lymphocytes to CNS tissue as a therapeutic target for stopping new lesion formation. High efficacy of B cell-depleting treatments showed that in MS, in contrast to EAE, B cells are essential for lesion development. This is likely linked to Epstein Barr virus (EBV) infection of B cells as a necessary, but insufficient trigger of MS (1). Because EBV does not infect rodents, EAE could not elucidate this crucial MS biology, showcasing the need for human studies.

Although the MS field benefitted greatly from visionary pathologists who performed highly informative observations (2–8), postmortem studies have understandable limitations. By studying CNS tissue once, usually in the final disease stage, pathology studies cannot differentiate disease consequences from its drivers. Additionally, pathology studies cannot examine CNS in its entirety and instead focus on a single disease aspect (e.g., MS lesions) in a limited number of subjects. Integrating such fractionated view into a system-wide understanding of the mutual relationships and the importance of mechanisms causing clinical disability is difficult. Finally, pathology provides this essential molecular information too late to inform therapeutic decisions.

Turning the impediment of relative inaccessibility of CNS into a scientific advantage can generate knowledge uniquely complementary to pathology studies. Specifically, by draining CNS interstitial fluid, the cerebrospinal fluid (CSF) contains analytes from all CNS cells, including infiltrating immune cells. CSF can provide longitudinal view of CNS tissue in health and disease. This study capitalizes on the ability of the National Institutes of Health (NIH) intramural research program to recruit patients willing to undergo research lumbar punctures (LP), and to deeply phenotype their disease using novel outcomes that, while enhancing sensitivity and accuracy, retain strong correlations with outcomes used by regulatory agencies for MS drug approval.

The study had two major goals: 1. To analyze measurements of thousands of CSF proteins and enumeration of CSF immune cells in MS and controls to identify pathways and upstream regulators that correlate with all measurable aspects of MS phenotype, including progression independent of relapse activity (PIRA). 2. To use machine learning to derive nine CSF biomarker-based models of aforementioned phenotypical aspects of MS that could predict imaging and clinical outcomes with robust statistical significance when applied to CSF samples derived from new cohort of MS patients. If this is possible, our last goal was to use the high correlation of novel outcomes with traditional MS outcomes to compare effect sizes of CSF biomarker-based models of Expanded Disability Status Scale (EDSS) (9), EDSS-based MS severity outcome Age-related MS severity Scale (ARMSS) (10) and traditional cognitive outcome, the Symbol digit modalities test (SDMT) (11) with the published literature.

## RESULTS

The study design is described in Figure1, Table 1, and Supplemental Table 1.

**Figure 1.**
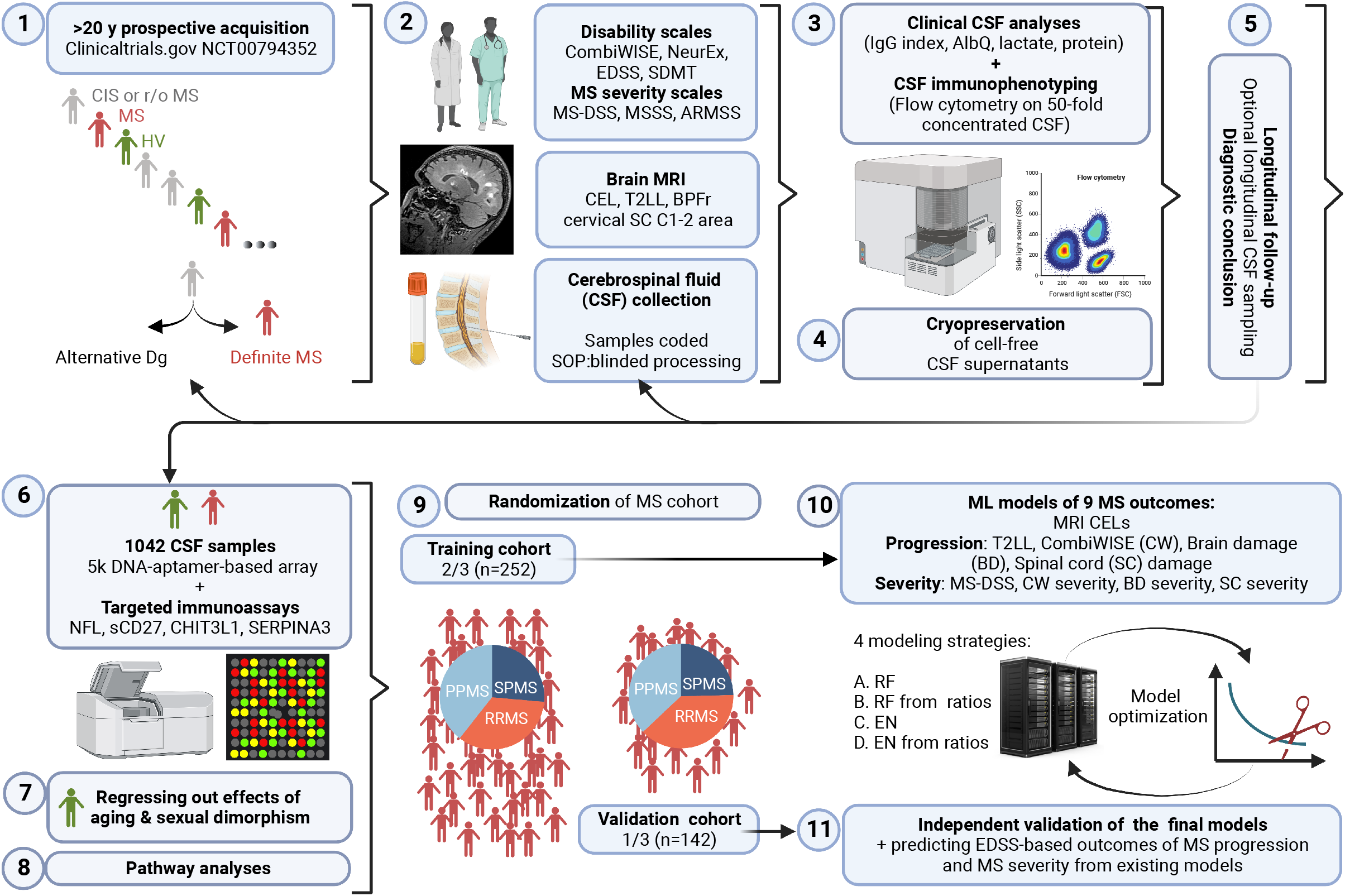
Study design. Patients with suspected or known multiple sclerosis (MS) diagnosis were enrolled into natural history protocol that collects standardized clinical, functional and brain imaging outcomes. Healthy volunteers (HV) undergo identical procedures. All subjects have cerebrospinal fluid (CSF) collection at protocol entry with optional follow-up LPs. Clinical CSF biomarkers are assessed by NIH clinical laboratory, while cellular CSF composition is analyzed by flow cytometry prospectively. CSF proteins were generated from cryopreserved coded samples by DNA-aptamer-based assay (SOMAScan), and in-house immunoassays (i.e., NFL, sCD27, CHIT3L1, and SERPINA3). CSF samples from HV were used to regress out effects of natural aging and sexual dimorphism on CSF proteins. Age- and sex-adjusted biomarkers were used for all downstream analyses. To generate nine CSF biomarker-based models of MS outcomes, the MS patients were randomized into training and validation cohort. In the training cohort, the outcomes were modeled using elastic net (EN) and random forest (RF) algorithms using single SOMAmers and SOMAmer ratios as predictors. An optimization pipeline on High Performance Computing cluster Biowulf generated final models. Appropriate models were recalibrated in the training cohort to predict widely used outcomes of MS disability (EDSS and SDMT) and severity (ARMSS). The models’ performance was evaluated in the independent validation cohort. Created with BioRender.com

**Table 1.** Demographic data.

|  | **HV** | **MS training** | **MS validation** |
| --- | --- | --- | --- |
| Number of samples | 71 | 647 | 324 |
| Number of subjects | 45 | 252 | 142 |
| Sex (% female) | 48.9 | 60.3 | 57.7 |
| Age at first LP (min-max) | 19.4 - 71.3 | 18 - 68.2 | 18.3 - 74.7 |
| Age at first LP (mean ± sd) | 37.7 ± 13.4 | 47 ± 11.9 | 47.4 ± 12.5 |
| Number samples/subject (min - max) | 1 - 4 | 1 - 8 | 1 - 7 |
| Number samples/subject (mean ± sd) | 1.6 ± 0.9 | 2.6 ± 1.7 | 2.3 ± 1.6 |
| years of follow-up (min - max) | 0 - 4.7 | 0 - 15.7 | 0 - 19.8 |
| years of follow-up (mean ± sd) | 0.9 ± 1.4 | 2.6 ± 3.3 | 2.4 ± 3.5 |

min - minimum, max - maximum, sd - standard deviation, LP - lumbar puncture, HV – healthy volunteers, MS – multiple sclerosis

### Development of comprehensive outcomes to measure different aspects of MS

Treatments that inhibit formation of MS lesions by >90% do not stop disability accumulation and are largely ineffective when started after age 54 (12). This suggests that different mechanisms mediate diverse aspects of MS, including PIRA. To investigate this hypothesis, we must first reliably measure diverse disease attributes.

Because individual genes and proteins exert small effect sizes on biological processes associated with complex (polygenic) diseases, success of biomarker modeling strategies strongly depends on the accuracy of modeling outcomes. The advantage of traditional MS outcomes, such as EDSS, is their broad use and easy quantification. The disadvantage is a lack of sensitivity (EDSS measures progression in a stepwise manner with an average patient progressing by 1 EDSS point per decade (13)), non-linear behavior, and weak signal to noise ratio (SNR; e.g., compare weak SNR for MRI volumetric outcomes (14) with strong SNR for contrast-enhancing lesions (CELs)). To transcend this problem, we developed and used novel outcomes with enhanced sensitivity, linearity, and SNR, while assuring that the novel outcomes retained strong correlation with traditional outcomes used for regulatory approval of MS drugs. This maximized the likelihood of obtaining reproducible insight from CSF biomarkers, while allowing verification that this insight is relevant for traditional outcomes by recalibrating models in the training cohort and predicting traditional outcomes in the independent validation cohort.

Using this strategy, we pursued comprehensive understanding of three main MS characteristics (Figure 2A): 1. *Formation of acute lesions* measured by (CELs) on brain magnetic resonance imaging (MRI). This process is strongly inhibited by MS treatments, peaks within years of MS onset, declining afterwards even in untreated patients (15). 2. *MS progression and cumulative CNS damage*: the cumulative volume of MS lesions (i.e., T2 lesion load; T2LL) increases from MS onset and peaks after CELs start declining, remaining relatively stable afterwards. This volumetric stability may be molecularly dynamic, where expansion of lesion edges might be compensated by the collapse of severely damaged lesion center. This hypothesis is supported by observations that loss of CNS tissue (measured as brain and SC atrophy) and accumulation of disability rise throughout MS duration, albeit at different rates for different subjects. 3. *MS severity*: these different rates of accumulation of CNS tissue destruction and irreversible disability reflect MS severity. Ideally, MS severity would be measured prospectively as patient-specific progression slopes. Practically, this is impossible because patients in natural history cohorts are treated with different medications and clinical trials are so short that only minority of patients experience sustained disability accumulation. Consequently, MS severity is measured by cross-sectional outcomes that relate MS progression to time, measured either as age or disease duration. Such cross-sectional MS severity outcomes effectively measure past rates of disability accumulation, by differentiating patients of the same age (or same disease duration) who accumulated more or less CNS damage (red-to-green triangles in Figure 2A). Because our goal was to measure MS progression and MS severity as comprehensibly as possible, we also pursued development of outcomes that differentiate brain from SC involvement.

**Figure 2.**
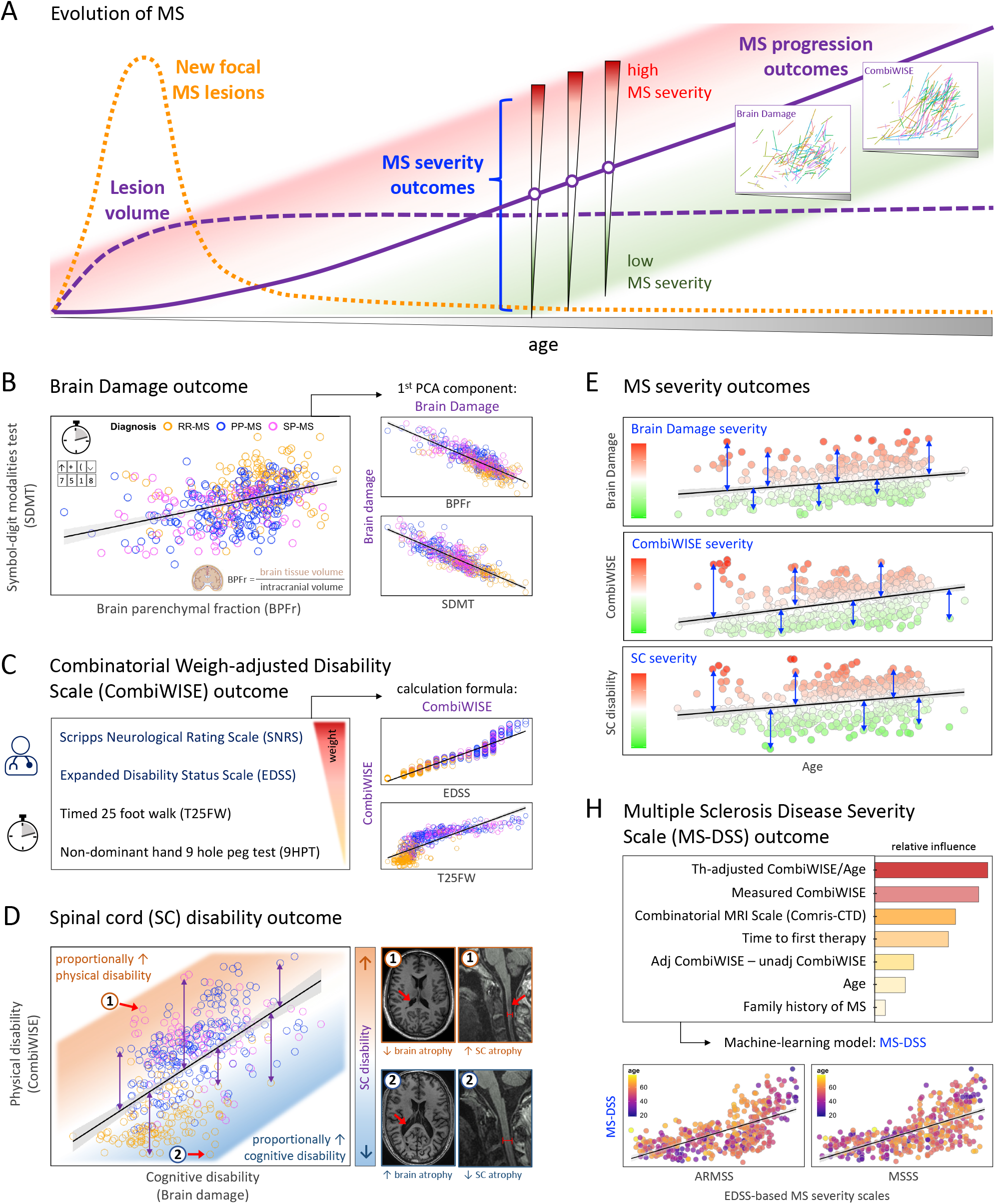
Development of novel, optimized outcomes. (**A**) Changes in MS disease characteristics over time. New MS lesions (yellow dotted line) start forming at MS onset and peak after several years, declining even in untreated patients. Each new lesion contributes to total lesion volume (purple dashed line) that remains relatively stable after focal lesions stop forming. The level of MS disability and CNS tissue destruction, collectively called: MS progression (purple solid line) increases steadily over the disease course. The inserts show MS patients longitudinal data for two MS progression outcomes brain damage [BD] and CombiWISE, demonstrating measurable linear progression slopes for both. Cross-sectional MS severity outcomes (blue curly bracket) differentiate subjects who accumulated more (red) from those who accumulated less (green) MS disability and CNS tissue damage in every age category. (**B**) BD outcome was generated as the first component of the principal component analysis of brain parenchymal fraction (BPFr derived from volumetric brain MRI analyses, x-axis) and cognitive test Symbol-digit modalities test (SDMT, y-axis). The BD outcome correlates strongly with its components (plots on the right). (**C**) Combinatorial Weight-adjusted Disability Scale (CombiWISE) is a linear combination of four disability scores (the red triangle shows decreasing weights of contributing scales), and it correlates strongly with traditional MS disability scales EDSS and T25FW (plots on the right). (**D**) Spinal cord (SC) disability outcome represents residuals of the linear regression model between CombiWISE (y-axis) and BD (x-axis). For subjects with comparable BD positive residuals (shades of orange) differentiate subjects who have proportionally more physical disability from those who have less physical disability (negative residuals; shades of blue). MRI images on the right illustrate examples of patients with (1) high SC disability (low level of brain atrophy and high level of SC atrophy), and (2) low SC disability (unremarkable cervical SC and high level of brain atrophy). (**E**) Three novel outcomes of MS severity were generated as age residuals of corresponding MS disability outcomes. The examples of residuals are depicted as blue double-arrowed lines, the levels of MS severity are shown in shades of green and red. (**H**) Multiple Sclerosis Disease Severity Scale (MS-DSS) is a machine learning-derived outcome that utilizes a combination of clinical, imaging, therapy, and demographic predictors, with their relative influence in MS-DSS represented by bar chart. MS-DSS correlates strongly with traditional, EDSS-based MS severity scales: ARMSS and MSSS.

Brain damage (BD) outcome was derived as the inverted first principal component of the correlation between normalized brain volume (brain parenchymal fraction; BPFr) and cognitive outcome SDMT (16) (Figure 2B). This limits noise in BPFr and SDMT measurements and yields outcome that progresses linearly in longitudinal testing (Figure 2A, BD insert) while retaining strong correlation with BPFr and SDMT (Pearson r −0.83 and −0.85, respectively, p-value < 2.2e-16).

Combinatorial Weight-adjusted Disability Score (CombiWISE) (14), a continuous scale from 0– 100, integrates four outcomes (Figure 2C) and measures linear disability accumulation (Figure 2A, CombiWISE insert). CombiWISE correlates strongly with EDSS (i.e., Spearman Rho 0.97, p-value < 2.2e-16); Figure 2C, right top panel and Figure 3A) and timed 25-foot walk (T25FW; Spearman Rho 0.88. p-value < 2.2e-16) (Figure 2C, right bottom panel).

**Figure 3.**
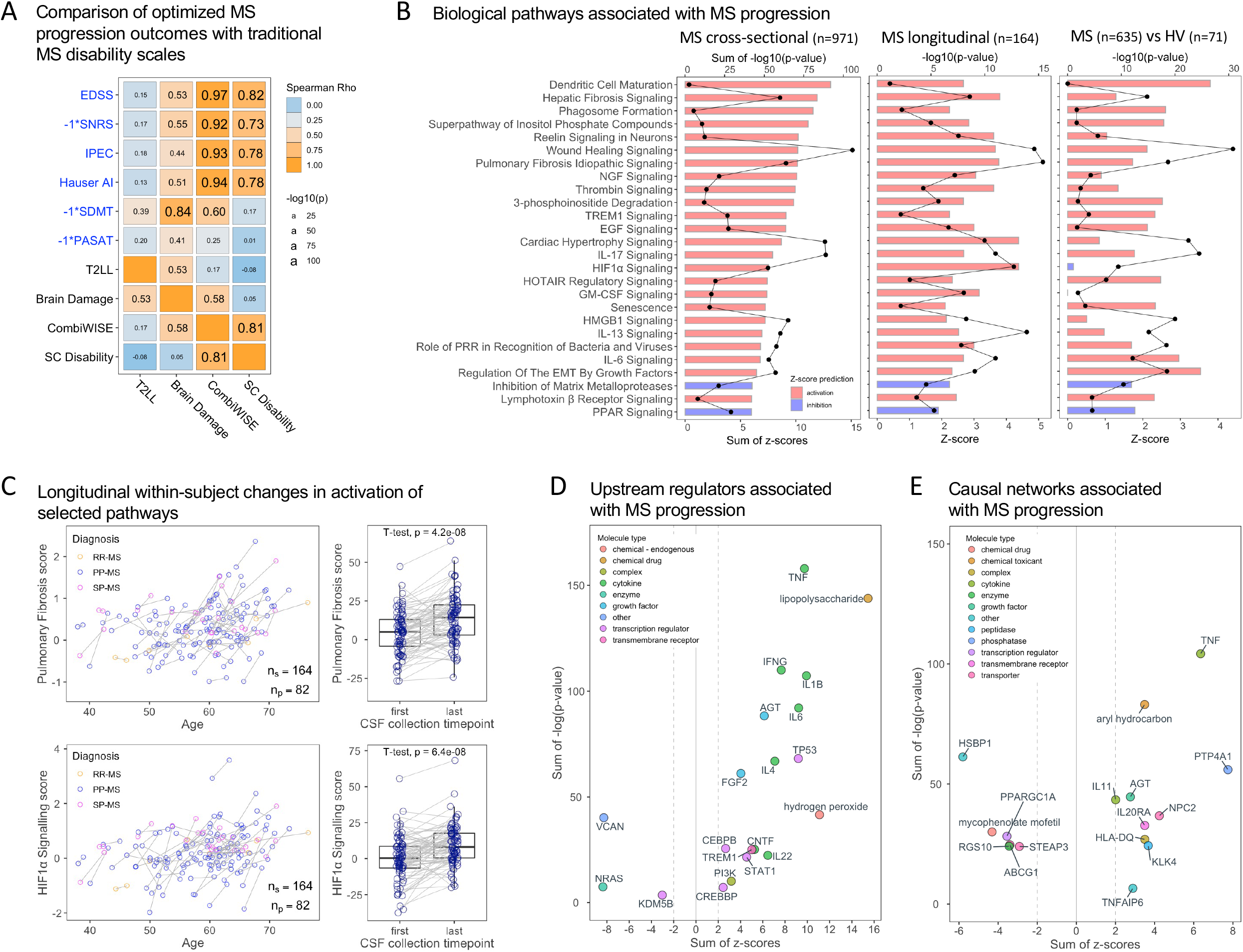
Biology of natural history of MS. (**A**) Correlation matrix between traditional MS progression outcomes (in blue) and optimized MS progression outcomes (in black). SNRS, SDMT, and PASAT scales (*) were inverted to retain the same directionality of all scores. Spearman correlation coefficients are displayed, their font size corresponds to −log_10_ *p*-value. (**B**) 26 canonical pathways were identified in the cross-sectional MS cohort (left plot) based on correlations with 4 MS progression outcomes. The effect sizes for individual pathways are shown as sum of *z*-scores horizonal bars, where red represent predicted activation and blue predicted inhibition of pathways with MS progression. The corresponding sum of −log_10_ *p*-value are depicted as connected black dots. Same pathways were significantly changed in longitudinal MS samples (middle plot; comparing first and last LP for each subject) and differentiated MS from HV (right plot) with identical directionality (**C**) Examples of longitudinal patient-level pathway scores (see Methods) of two predicted activated pathways, pulmonary fibrosis (top plots) and HIF1α signaling (bottom plots), showing statistically significant increase over time. (**D**) A selection of upstream regulators and (**E**) causal networks associated with MS progression are displayed with sum of *z*-score across the four progression outcomes on x-axis and sum of −log_10_ *p*-value on the y-axis. The color of each point demonstrates the molecule type for identified features.

To isolate SC disability, we used residuals of linear regression of BD and CombiWISE (Figure 2D). For any given BD value, subjects with high SC disability will have accumulated more physical disability than subject with low SC disability. SC disability differentiates subjects with low brain and high SC damage (Figure 2D, MRI inserts for patient 1) from those with high brain and low SC involvement (MRI inserts for patient 2).

Collectively, T2LL, BD, CombiWISE, and SC disability will allow identification of molecular processes associated with all types of MS progression. We hypothesized that modeling CSF biomarkers against these outcomes will validate CNS processes seen in MS autopsy studies. However, this analysis will not differentiate processes induced by MS from those that may drive CNS destruction.

Studying MS severity increases the probability of identifying such causal processes, as it focuses on mechanisms that differentiate slow from fast progressors among subjects of all ages and disability levels. We used residuals of linear regression models between age and three MS progression outcomes (i.e., BD, CombiWISE, and SC disability; Figure 2E) to derive cross-sectional severity outcomes that capture both cognitive and physical disability. We also included MS Disease Severity Scale (MS-DSS (17)), which integrates clinical and MRI outcomes and adjusts for effects of treatments (Figure 2H). MS-DSS correlates with traditional, EDSS-based MS severity outcomes, MS Severity Scale (MSSS; Spearman Rho 0.68, p-value < 2.2e-16), and ARMSS (Spearman Rho 0.66, p-value <2.2e-16) (Figure 2H, lower panels). For reference, modeling outcomes are also summarized in Table 2.

**Table 2.** Summary of MS progression and MS severity outcomes used in the study.

| Outcome | Acronym | MS process | Description | Reference |
| --- | --- | --- | --- | --- |
| T2 lesion load | T2LL | Progression | Volume of T2 lesions generated from T1- and T2-weighted images using lesionTOADS algorithm integrated into Qmenta platform | (57) |
| Brain damage | BD | Progression | First component of the principal component analysis of brain parenchymal fraction (measured as percentage of brain parenchyma volume of the skull volume) and SDMT (measuring cognitive function) | This study |
| Combinatorial Weight-adjusted Disability Score | CombiWISE | Progression | Machine learning model that combines weighted disability scores: EDSS, SNRS, T25FW, and nondominant hand of the 9HPT | (14) |
| Spinal cord disability | SC disability | Progression | Residuals of the linear regression model between BD (measuring cognitive disability) and CombiWISE (measuring physical disability) | This study |
| Brain damage severity | BD severity | Severity | Age residuals of BD | This study |
| CombiWISE severity | CombiWISE severity | Severity | Age residuals of CombiWISE | This study |
| Spinal cord severity | SC severity | Severity | Age residuals of SC disability | This study |
| MS Disease Severity Scale | MS-DSS | Severity | Machine learning model that combines measured disability, therapy intervention, age, and MRI grading of the CNS destruction | (17) |

### Molecular mechanisms associated with natural history of MS progression

Correlation matrix (Figure 3A) relates these new, optimized MS progression outcomes to traditional scales of cognitive (i.e., SDMT and Paced Auditory Serial Addition Test [PASAT] (18)) and physical disability (EDSS, Scripps Neurological Rating Scale [SNRS] (19), Instituto de Pesquisa Clinica Evandro Chagas [IPEC] scale (20), and Ambulation Index [AI] (21)). Strong positive correlations demonstrate that CombiWISE successfully captures all traditional measures of physical disability. Observing that BD correlates the strongest with cognitive disability outcomes and brain T2LL, while SC disability does not correlate with cognitive disability outcomes, T2LL or BD, confirmed achievement of stated goal of separating brain from SC damage.

Next, to isolate MS-associated biology, we regressed out effects of physiological aging and sex on CSF proteins measured by DNA-aptamers (i.e., SOMAmers) using longitudinal healthy volunteers (HV) CSF data. All subsequent analyses use HV age/sex-adjusted biomarkers.

To uncover the biology of MS progression, we identified biomarkers that correlated significantly (after false discovery rate [FDR]-adjustment of *p*-values) with MS progression outcomes (Supplemental Table 2) and uploaded their correlation coefficients into Ingenuity Pathway Analysis (IPA^®^). IPA^®^ contains over 8.5 million instances of manually curated knowledge about relationships between transcripts, proteins and molecular pathways. Figure 3B, left panel contains sum of *z*-scores and sum of −log_10_ *p*-values for pathways significantly associated with at least two progression outcomes. Most pathways have positive *z*-scores (shown in red), indicating their predicted activation during MS progression. Majority are neuroinflammation-related, such as IL17, GM-CSF, IL13, LTb, and IL6 signaling, dendritic cell (DC) maturation, high mobility group box protein 1 (HMGB1) signaling, Triggering Receptor Expressed on Myeloid Cells 1 (TREM1) signaling, phagosome formation, and role of pattern recognition receptors (PRR) in recognition of bacteria and viruses. However, we also identified strong fibrosis signature: hepatic and pulmonary fibrosis, wound healing, cardiac hypertrophy, and regulation of epithelial-mesenchymal transition (EMT) by growth factors such as epidermal growth factor, and by long noncoding RNA (lncRNA) HOTAIR. Hypoxia, reflected by activation of hypoxia inducible factor 1 subunit alpha (HIF1α), likewise induces EMT. Senescence and thrombin signaling pathways were also activated with MS progression. Finally, neuronal reelin signaling and neurotrophic growth factor (NGF) signaling induced by MS progression likely reflect response of CNS tissue to injury. Two pathways (inhibition of matrix metalloproteinases and PPAR signaling) had negative *z*-scores, indicating their predicted suppression during MS progression.

Our observation that neuroinflammation *increases* during MS progression contradicts the prevailing belief that neuroinflammation *decreases* with MS progression. Therefore, we employed longitudinal samples from untreated MS patients (collected mostly during placebo-controlled secondary progressive MS [SPMS] (22) and primary progressive MS [PPMS] (23) clinical trials) to investigate intra-individual changes of these pathways during MS progression. Because none of these patients experienced exacerbations during this longitudinal follow-up, this cohort allows us to study intrathecal processes that correlate with PIRA.

This analysis was consistent with the cross-sectional cohort (Figure 3B, middle panel), even though longitudinal data consisted of pathways’-specific scores at first and last untreated CSFs (see Methods). Figure 3C exemplifies individual longitudinal data for two of these pathways (i.e., pulmonary fibrosis and HIF1α signaling). Finally, we observed that activation of these pathways also differentiates MS from HV (Figure 3B, last panel).

To expand insight into regulators of identified pathways, we leveraged the ability of IPA^®^ to integrate CSF biomarkers into predictions of upstream regulators (Figure 3D; molecules directly upstream of measured biomarkers) and causal networks (Figure 3E; molecules several levels upstream of measured biomarkers) that may operate in meninges and CNS parenchyma. This analysis strengthened the conclusion that neuroinflammation increases with MS progression, highlighting the role of TNF, IFNγ, IL1β, IL4, IL6, IL11, IL20, IL22, and ligands such as lipopolysaccharide (LPS), aryl hydrocarbon (previously linked to MS and EAE (24)), and hydrogen peroxide generated during oxidative stress.

### Predominant involvement of brain versus SC in MS is linked to different biology

Developing outcomes that separated cognitive disability linked to BD from physical disability originating from SC allowed search for biological underpinnings of these topological differences. Propensity score matching (Figure 4A) identified MS patients with comparable BD, but different physical (CombiWISE) disability. Because SC disability outcome is novel, we formally tested whether it reflects MS-associated SC injury by correlating it with the cross-sectional SC imaging biomarker measured as C1-2 SC area (Figure 4B). Significant negative correlation proved that BD-CombiWISE residuals reflect SC damage and that propensity score-matched groups have vastly different SC atrophy.

**Figure 4.**
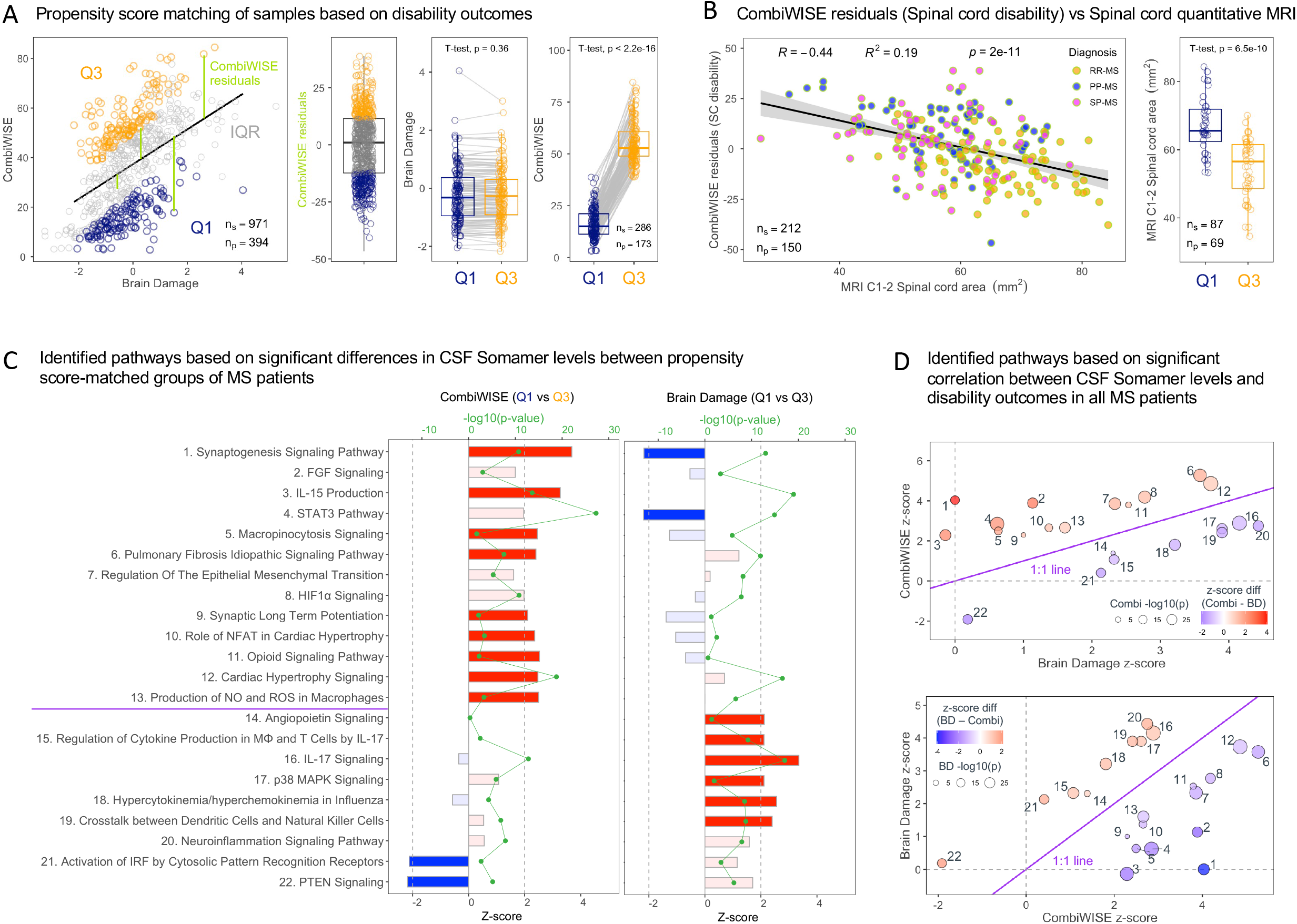
Biology of MS-related brain vs spinal cord damage. (**A**) To separate MS patients with predominant brain versus (SC) disease, we calculated SC disability as the residuals (green lines) of the linear regression model between BD and CombiWISE. Residuals within interquartile range (IQR, gray points) were removed, and the groups of patients with residuals below the 1^st^ quartile (Q1, dark blue points) and above the 3^rd^ quartile (Q3, orange points) were paired using propensity score matching on BD outcome. (**B**) Left plot: To prove that SC disability reflects SC damage, we correlated it (y-axis) with upper cervical SC area measured by MRI at C1-C2 level (x-axis) in the MS cohort. Right plot: MS patients with low SC disability (Q1; dark blue) had significantly greater cervical SC volume than patients with high SC disability (Q3; orange). (**C**) Statistically significant differences between Q1 and Q3 patients identified 22 pathways significantly activated (red horizontal bars) or inhibited (blue horizontal bars) when comparing patients with predominant SC (left plot) versus brain (right plot) involvement. Horizontal bars with light blue or light red color didn’t reach the significant *z*-score cut-off (depicted as vertical dashed lines). The FDR-adjusted −log_10_ *p*-values for all pathways are illustrated by green connected dots. (**D**) In sensitivity analyses the 22 pathways discovered in panel **C** were also identified by comparing BD *z*-scores (x-axis) and CombiWISE *z*-scores (y-axis). Top plot shows that pathways 1–13 activated (above the purple 1:1 line) in patients with proportionally increased CombiWISE. Conversely, pathways 14–22 were activated in MS patients with proportionally higher BD outcome (bottom plot).

We used these matched MS patients to identify pathways activated/inhibited in subjects with comparable BD but different SC disability (Figure 4C, left panel) or, analogously, in subjects with comparable SC disability but different BD (Figure 4C, right panel). Both comparisons yielded consistent results: MS patients with proportionally higher SC disability had activated pathways related to fibrosis (FGF signaling, STAT3 pathway, pulmonary fibrosis, the regulation of EMT, HIF1α, and cardiac hypertrophy); and to CNS response to injury (synaptogenesis, synaptic long-term potentiation). In contrast, patients with proportionally higher BD showed activation of neuroinflammation pathways, especially IL17, p38 MAPK, and innate immune cells such as DCs, natural killer (NK) cells, and macrophages. Pathways activated in one phenotype were generally repressed in the other.

As sensitivity analyses, we used all MS subjects to assess significant correlations between CSF biomarkers and the two divergent progression outcomes. The same pathways (Figure 4D) were associated with predominance of BD versus CombiWISE with high statistical significance.

### Biological mechanisms with stronger association with MS severity than MS progression

Mechanisms induced by MS (i.e., epiphenomena) would be underrepresented at the beginning of MS and overrepresented in older people with longer disease duration, who accumulated more disability. In other words, processes we identified as correlating with MS progression may represent such epiphenomena. Some of these MS-induced mechanisms may still be causal, meaning that once expressed, they may contribute to CNS injury. Such causal processes should, in addition to correlating with MS progression outcomes also correlate with MS severity outcomes.

Yet another category of causal mechanisms may drive CNS injury from the earliest stages of MS through entire MS duration: these processes would differentiate fast progressing from slow progressing MS patients across all age/disability levels. In other words, these candidate pathogenic processes would correlate with MS severity outcomes, but not with MS progression outcomes. Thus, to identify candidate disease mechanisms, we searched for biological processes reflected by CSF biomarkers with much stronger correlation with MS severity than MS progression outcomes.

Because the MS severity outcomes employed here are novel, we first assessed their correlation with traditional EDSS-based MS severity outcomes (Figure 5A). CombiWISE severity best reflects EDSS-based severity outcomes, achieving especially strong correlation with ARMSS (i.e., Spearman Rho 0.96, R^2^ 0.86, p-value < 2.2e-16). Weaker correlations of BD severity with MSSS (Spearman Rho 0.46, R^2^ 0.22, p-value 7.6e-12) and ARMSS (Spearman Rho 0.50, R^2^ 0.24, p-value 7.5e-14) reflects poor sensitivity of EDSS for cognitive disability and highlights added value of BD severity to identify patients with different rates of accumulation of cognitive disability.

**Figure 5.**
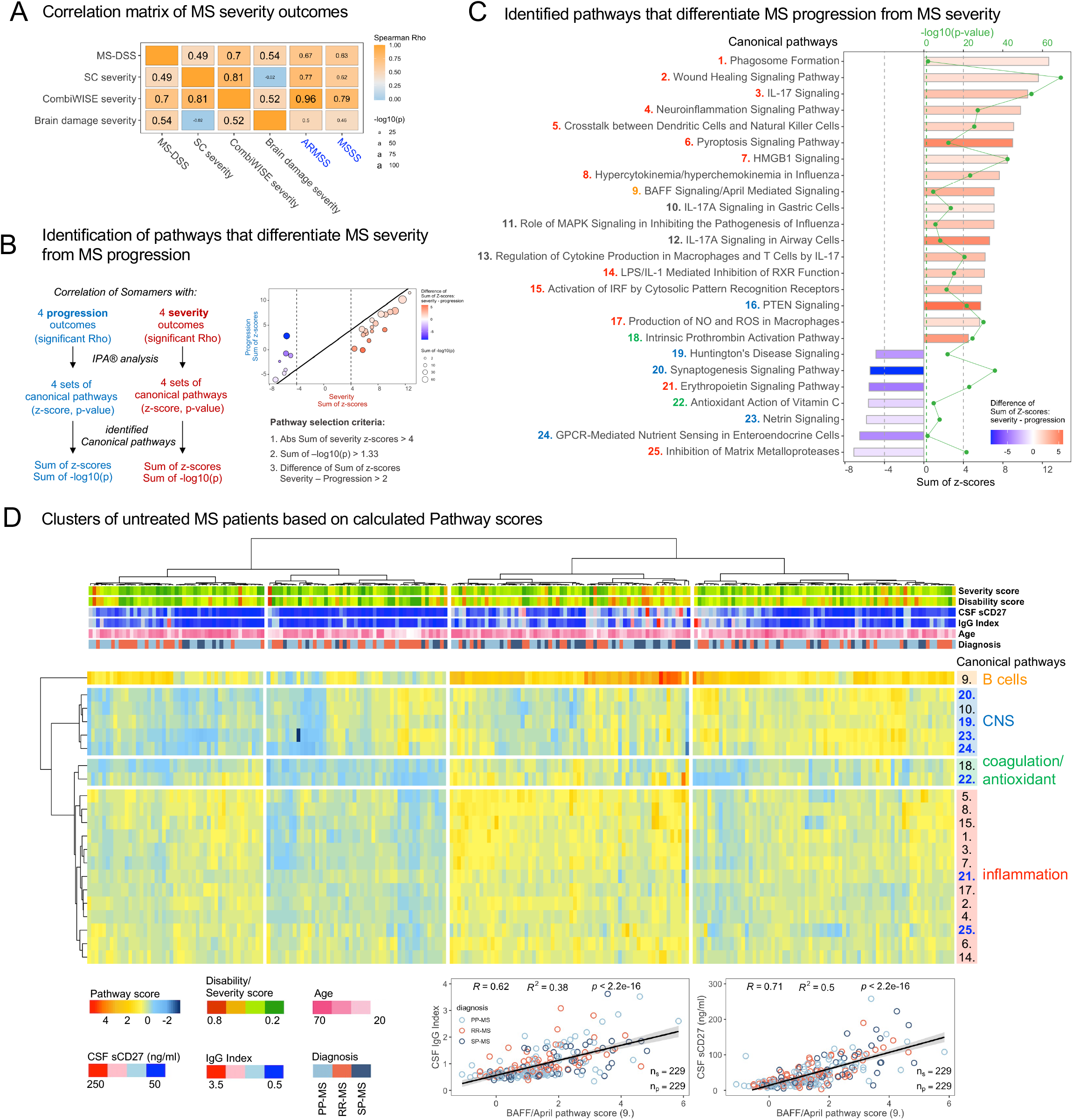
Biology of MS severity. (**A**) Correlation matrix of traditional EDSS-based MS severity outcomes (ARMSS and MSSS) and optimized MS severity outcomes used in this study. Spearman correlation coefficients are displayed, the font size corresponds to the −log_10_(*p*-value). (**B**) Flowchart of pathway identification: IPA-identified pathways for four progression outcomes (blue) and four severity outcomes (red) were merged and the sum of *z*-scores and sum of −log_10_ *p*-value were calculated for each pathway. Pathways that differentiate MS severity from MS progression had to fulfill the three displayed pathway selection criteria. (**C**) 25 pathways more strongly (and significantly) associated with MS severity than MS progression outcomes. The sum of *z*-scores is displayed as length of the horizontal bar (positive for activation, negative for inhibition), the shades of blue and red fill illustrate difference between sum of *z*-scores for severity and progression outcomes. Sum of −log_10_ *p*-value is displayed as green connected dots. (**D**) We calculated patient-specific activation scores for selected pathways (see Methods) for untreated MS patients at first LP. Unsupervised algorithm clustered patients (columns) into 4 clusters, and 25 pathways from (**C**) into 4 clusters (lines). Patient clusters differed in the level of activation of B cell/plasma cell-activation pathway (orange), CNS cluster (blue), coagulation/antioxidant cluster (green), and inflammation cluster (red). The numbers and colors of individual pathways are identical in panels **C** and **D**. Additional outcomes/biomarkers are displayed above the heatmap, illustrating distribution of age, diagnoses, disability, and severity outcomes, as well as broadly accepted biomarkers of intrathecal inflammation sCD27 and IgG index. Two plots on the bottom illustrate strong correlation between activation score for pathway 9 – BAFF/APRIL signaling and two biomarkers of intrathecal inflammation – IgG Index (left) and CSF levels of sCD27 (right).

Next, using the stated rationale we searched for pathways that correlated with multiple MS severity outcomes, while simultaneously correlating stronger with MS severity than MS progression (Figure 5B and Methods). These pathways are shown in Figure 5C, while upstream regulators and causal networks are in Supplemental Table 3.

Although the neuroinflammation-related pathways also dominated this analysis, we observed that some selected pathways were new (i.e., not associated with MS progression), such as activation of B cells and plasmablasts/plasma cells by the B cell activation factor [BAFF] and a proliferation-inducing ligand [APRIL] and pathways related to viral infections (i.e., activation of IRF by cytosolic PRRs, influenza-related pathways and pyroptosis). On the side of innate immunity, the new pathways associated with MS severity but not MS progression were production of nitric oxide (NO) and reactive oxygen species (ROS) in macrophages, crosstalk between dendritic cells (DC) and natural killer (NK) cells and lipopolysaccharide (LPS)/IL-1 mediated inhibition of retinoid X receptor (RXR).

Other neuroinflammatory pathways, such as those linked to Th17 immunity, HMGB1 signaling and Inhibition of Matrix Metalloproteinases (the latter with negative z-score, which predicts beneficial effect on MS severity) represent examples of pathways that were associated with MS progression but passed this analysis as well due to their stronger correlation with MS severity outcomes.

Finally, we note that majority of fibrosis-related pathways (except for related wound healing signaling pathway) did not pass our stringent criteria and thus we consider them only associated with MS progression, but not MS severity. We also failed to identify any inflammation-unrelated true neurodegenerative mechanisms in this analysis. In fact, CNS pathways we identified, such as synaptogenesis and netrin signaling had negative z-score, indicating that they are over-represented in MS patients who progress slower. Analogously, Erythropoietin signaling, and Antioxidant action of vitamin C were predicted as beneficial pathways.

### Intra-individual heterogeneity and multiplicity of pathways linked to MS severity

Unsupervised clustering of MS severity-related pathways (Figure 5D) separated MS patients into four clusters. Inhibition of matrix metalloproteinases and Erythropoietin signaling, two of the seven “beneficial” pathways for MS severity (blue font in Figure 5D), clustered with neuroinflammation, consistent with their immunoregulatory role (25). Remaining beneficial pathways of compensatory response of CNS tissue clustered together. Age, sex and traditional MS categorization did not influence clustering, confirming that most of the identified candidate causal pathways are common to all ages and disability levels.

Because IPA^®^ aggregates knowledge from diverse experimental systems and because the relative concentrations of CSF biomarkers were heavily processed, we asked whether these complex analyses reflect tangible processes. Therefore, we assessed correlations between patient-specific pathway activation scores (see Methods) and previously validated biomarkers of MS-related CNS inflammation measured by different assays.

Broadly used neuroinflammation biomarkers such as IgG index and sCD27 demonstrated strong correlation with patient-specific activation scores of inflammatory pathways: Figure 5D, lower panels show their correlation with BAFF/APRIL signaling pathway, representing the strongest correlations observed. Additional weaker, but still significant correlations are shown in Supplemental Figure 1. Pyroptosis was the MS severity pathway with the broadest association with previously identified MS biomarkers, including CHI3L1 and SERPINA3.

Integrating prospectively acquired flow-cytometry based CSF cellular data with proteomic analysis shows that neuroinflammation associated with MS severity and MS progression is compartmentalized to CNS tissue and meninges, due to the simultaneous decrease in absolute number of B cells, T cells, and NK cells in the CSF of MS patients associated with MS progression (Supplemental Figure 2). This interpretation is supported by observations that no cellular CSF biomarkers correlated with MS progression or severity outcomes (not shown).

### CSF biomarkers can be integrated into predictive models of clinical and imaging aspects of MS

Our final goal was to assemble CSF biomarkers into models aimed to predict all 9 MS outcomes and to compare effect sizes of such models with the published literature.

To gain insight into the nature of relationships between CSF biomarkers (i.e., linear versus non-linear) we used different modeling strategies (Figure 6A): the multiple linear regression represented by elastic net (EN) models and random forest (RF) representing tree-based algorithms. Additionally, being concerned that our training cohort could be too small in relationship to the number of predictors, we compared the models from 5,034 SOMAmers with models from 12.9 million SOMAmer ratios. As the models based on SOMAmer ratios had >2500-fold higher numbers of predictors per subject, if our training cohort was of inadequate size for ML algorithms to find optimal solutions, the SOMAmer ratio-based models should have higher degree of overfit, leading to weaker validation.

**Figure 6.**
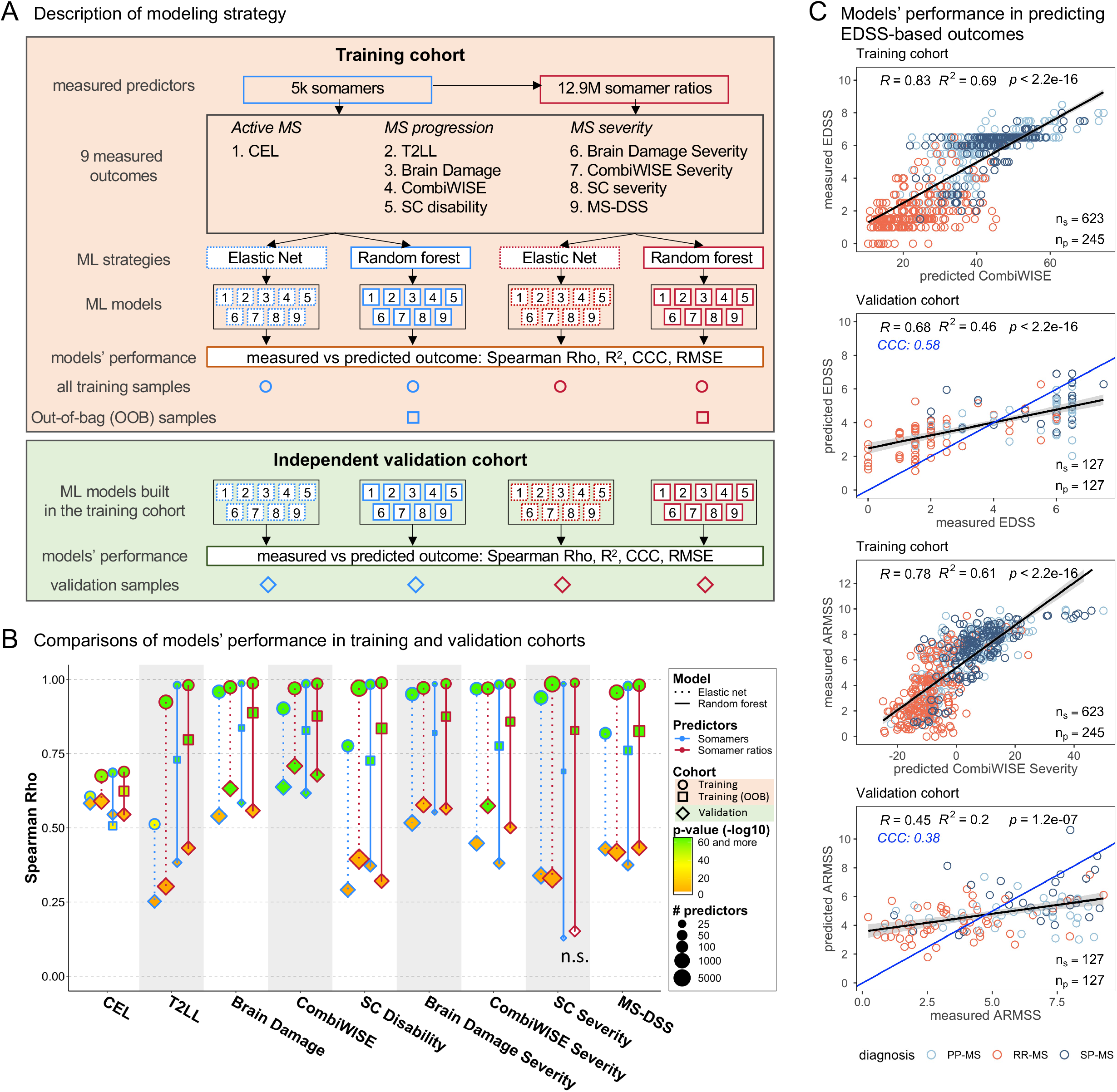
CSF biomarker-based models of MS outcomes. (**A**) In the training cohort (orange box), two parallel modeling approaches were followed – using 5,034 SOMAmers (blue) or 12.9 million of SOMAmer ratios (red) as predictors to model 9 already introduced MS outcomes. Each outcome was modeled using two algorithms – elastic net (EN) linear regression and tree-based algorithm random forest (RF). For RF models, out-of-bag (OOB) predictions were evaluated as more realistic estimate of effect sizes from training cohort data. The generated models were then tested in the independent validation cohort (green box) that did not contribute, in any way, to model generation. (**B**) A comparison of models’ performance based on Spearman Rho in training (circles), OOB (squares), and validation (diamonds) cohorts. The EN models are illustrated by dotted lines connecting training and validation cohort sets, while RF models are connected with solid lines. Models built with single SOMAmers as predictors are in blue, models generated from SOMAmer ratios are shown in red. The size of the points represents the number of predictors retained in the final model, the fill of the datapoints illustrates −log_10_ *p*-value. Two RF models of SC severity did not validate (n.s.). (**C**) To compare effect sizes of CSF-based models with published literature, we recalibrated appropriate models (i.e., CombiWISE for EDSS and CombiWISE severity for ARMSS) based on strong correlations between CombiWISE and EDSS and CombiWISE severity and ARMSS in the MS training cohort. Specifically, SOMAmer-predicted CombiWISE was used to generate a simple linear regression model of measured EDSS in the training cohort (top plot). The regression equation was then used in the validation cohort to predict EDSS (second plot from the top). The blue line represents 1:1 line. Analogous method was used for predicting ARMSS (bottom two plots).

In the training cohort (Figure 6B shown as circles; Supplemental Table 4): the RF models outperformed EN models, even though EN models included more biomarkers (thousands) compared to RF models (up to hundreds). This suggested non-linear relationships in the structure of CSF biomarkers and pathways they represent. Additionally, models from biomarker ratios outperformed models from single SOMAmers. The out-of-bag (OOB) RF results are often used in lieu of validation strategy, as they reflect model’s performance on the samples that the algorithm omitted from the specific iteration(s) of the model. Consistently, OOB results (Figure 6B; squares) showed diminished effect sizes for all models.

However, the OOB results, like all cross-validation strategies that reuse training cohort, contain circular argument (26): these samples did contribute to some aspects of the final models, e.g., the selection of predictors. Therefore, the true effect sizes can be derived only from a new cohort of patients, whose samples did not participate in any aspects of model development (27).

When final models were applied to independent validation cohort (Figure 6B diamond shapes), it became obvious that CSF proteins, like previously shown for genes, exert mostly linear (additive/subtractive) effects on clinical outcomes, as EN models validated with comparable or higher effect sizes than RF models (e.g., see SC severity). The models from SOMAmer ratios also validated comparable or higher effect sizes to models from individual SOMAmers, proving that our training cohort was sufficiently large to find optimal solutions.

Two observations showcase the necessity to use independent validation for assessing ML-based models: first the effect sizes for all models decreased significantly when compared to OOB data. Second, the OOB data did not predict even the hierarchy of model validation: e.g., the RF SC severity models did not validate, even though their OOB performance strongly outperformed CEL models, which validated with some of the highest effect sizes (Figure 6B; Supplemental Figure 3).

The most insightful result from comparing the different modeling strategies is that *the strongest determinant of validated effect sizes was the modeling outcome*. While we validated models for all MS outcomes with very low *p*-values, the hierarchy of effect sizes strongly suggest that the limiting factor in model’s performance is the outcome: the level of accuracy with which we can measure it and the degree to which it reflects tangible biological processes.

### CSF-biomarker based models reliably predict traditional MS outcomes in the independent validation cohort

Although the hierarchy of validated effect sizes support our premise that we need accurate outcomes to successfully model different MS characteristics; the major drawback of these optimized outcomes is that they are not used broadly. However, based on strong correlations between novel outcomes and traditional MS outcomes (Figures 1–3), we knew we can mathematically recalibrate CSF-biomarker models in training cohort to predict traditional outcomes (Figure 6C).

We used CSF-biomarker predicted CombiWISE to derive linear regression model with measured EDSS (Figure 6C, top panel). Applying this correction to the validation cohort (Figure 6C, second panel from the top), we observed correlations between measured and CSF-biomarker predicted EDSS that explained 46% of variance (p-value <2.2e-16) with concordance correlation coefficient (CCC)=0.58 (i.e., CCC = 1 represents exact concordance between measured and predicted outcomes). Analogously, predicted ARMSS (Figure 6C, lower two plots) in the independent cohort explained 20% of variance (p-value 1.2e-7) with CCC=0.38. Finally, we used CSF-predicted BD to derive a linear model with measured SDMT. The model-predicted SDMT in the validation cohort explained 36% of variance (p-value 1.1e-11) of the measured SDMT with CCC= 0.54 (Supplemental Figure 6).

Based on the meta-analysis of MS models (28), these are the strongest validated effect sizes for these 3 traditional outcomes used for regulatory approval of MS drugs, using any type of predictors (i.e., clinical/demographic, MRI, blood/CSF biomarkers or genes).

## DISCUSSION

Mechanistic understanding of the MS lesions and their reliable quantification by CELs spearheaded development of treatments that virtually eliminate their formation. This study elucidates processes associated with natural history of MS beyond the formation of MS lesions, including PIRA, which represents currently the most pressing therapeutic need in MS.

Because it is impossible to discuss every molecule in this rich dataset, Supplementary Tables contain comprehensive analyses of single protein(s) with all raw data acquired in this study. While we reproduced most studies linking specific protein(s) with MS, single proteins exerted (predictably) much smaller effect sizes on MS outcomes compared to pathways that aggregate proteins based on their biological relationships. Therefore, we focused the results, and we’ll focus the discussion on these pathways.

The most surprising result of current study is the dominance of neuroinflammation-related pathways among biological processes that correlate with both MS progression and MS severity. As current MS drugs inhibit inflammation and formation of inflammatory CELs, their decreasing efficacy in later stages of MS, characterized by PIRA, led to broad belief that progressive MS (PMS) is *less* inflammatory than relapsing-remitting MS (RRMS) and that neurodegenerative mechanisms drive disability accumulation in PMS (29). In contrast, our results show that intrathecal activation of (some) neuroinflammation-related pathways *increases* with MS progression. This was based on the analysis of CSF biomarkers that correlate with diverse MS progression outcomes and validated by biomarkers that significantly changed in longitudinal CSF samples from placebo arms of PMS clinical trials. Because none of these progressive patients experienced MS relapse during this longitudinal follow-up, they represent PIRA cohort.

The second unexpected observation was that our analyses did not identify any inflammation-unrelated neurodegenerative pathways that would positively correlate with MS progression or MS severity. This lack of inflammation-unrelated neurodegenerative mechanisms validates genotyping data that linked expression of genes associated with MS susceptibility alleles to the cells of the immune system (including its intrathecal components such as microglia and astrocytes), but not neurons (30).

The third unexpected observation was that inflammatory pathways that correlate with MS progression did not comprise one unique phenotype: they included both innate and adaptive immunity of Th17 (IL17, GM-CSF and IL6), Th1 (IFNγ and TNFα) and Th2 (IL13 and IL4) phenotype. The notable exception was lack of pathways associated with humoral adaptive responses represented by B cells, plasma cells/plasma blasts and antibodies, which are hallmark of MS.

While Th2 T cells are beneficial in acute experimental allergic encephalomyelitis models of MS, Th2 inflammation may also cause fibrosis, which we found strongly linked to MS progression. IL11, another upstream regulator identified in our analyses, is almost exclusively expressed in fibroblasts and linked to many fibrotic conditions (31).

Fibrosis is associated with EMT, a process in which epithelial cells lose their tight-junctions/desmosome-mediated association with basal membranes and with each other. Dislodged epithelia differentiate into myofibroblasts, gaining invasive properties. Many signaling pathways, e.g., eGFR, WNT/beta-catenin, NOTCH, PI3K/AKT, NFkB, and hypoxia, induce EMT. We show that activation of upstream regulators of these pathways increases with MS progression. Although fibrosis is an end result of many chronic inflammatory processes, literature search demonstrates that its intrathecal activation has not been previously recognized in MS. If novel anti-fibrotic agents cross BBB, they might be tested as adjunctive treatments for (some) MS patients, although we acknowledge that fibrosis was more strongly associated with MS progression than MS severity, suggesting that it may represent an epiphenomenon.

Another novel finding is that different biological processes underlie predominant involvement of brain versus SC by MS: neuroinflammation was associated with predominant involvement of brain and fibrosis with SC-predominant MS. Of course, we cannot determine which drives which: whether CNS location modulates phenotype and outcome of the inflammatory infiltrate, or whether propensity to different phenotype/outcome of inflammation determines location of CNS injury. Only future studies, ideally linked to genetics, might answer this emerging question.

Perhaps the most important finding, with direct relevance for future drug development, is our identification of pathways that correlate with MS severity. While neuroinflammatory processes dominated this analysis as well, there were some notable differences that may provide insight into MS disease mechanisms. We mentioned previously that pathways linked to activation of the humoral arm of adaptive immunity were not associated with MS progression but were associated with MS severity. Indeed, these pathways correlated with IgG index, which is elevated in vast majority of MS patients from the onset of MS till its end. Likewise, while biomarkers released by all T cell phenotypes were linked to MS progression, Th17 pathways were more strongly associated with MS severity than MS progression. Equally, activation of innate immunity was linked to both MS progression and MS severity, but pathways linked to MS severity were enriched for effector mechanisms that mediate tissue damage, such as NK cells, generation of NO and ROS and pyroptosis. Three pathways related to viral infections were also exclusively associated with MS severity, but not MS progression.

If we were to integrate the gained knowledge into a single hypothesis, it would be that EBV reactivation of latently infected B cells, perhaps with lytic infection of some CNS epithelial cells, not only triggers MS, but continues fueling the compartmentalized inflammation. Resulting pyroptosis, identified in MS lesions (32), may be an important mechanism of CNS cells death in MS. Indeed, MS-derived EBV-infected CSF B cells have higher lymphangiogenic potential compared to controls (33) and therefore may contribute to formation of tertiary lymphoid follicles, a hallmark of compartmentalized inflammation in MS (34). However, EBV presence in MS CNS remains controversial (35, 36), requiring further research to investigate the hypothesized link between EBV and MS progression.

Current study has following limitations: 1. The IPA knowledge base and therefore our pathway analyses integrate knowledge from varied sources such as human and animal in-vitro and in-vivo data derived from different biological fluids or organs, with human CNS being likely under-represented. One may wonder how relevant our conclusions are to human CNS tissue. Reassuringly, processes our pipeline linked to MS progression overlap greatly with processes seen in postmortem MS CNS, such as hypoxia, activation of clotting cascade, compartmentalized inflammation, and microglia/macrophage activation with generation of NO and ROS (4, 37–39). Additionally, patient-specific pathway scores we generated using bioinformatics pipeline correlated highly with relevant validated biomarkers: e.g. BAFF/APRIL signaling pathway correlating with IgG index. Nevertheless, we acknowledge that some recently discovered biological processes linked to MS severity, such as toxic astrogliosis (40–42), are not yet annotated in IPA and were missed. 2. Although subtracting effects of physiological aging and sexual dimorphism on CSF biomarkers helped to identify MS-related processes, it likely under-estimated effect sizes with which CSF biomarkers can predict MS progression and severity outcomes. This is because both age and sex affect MS progression and MS severity. 3. We did not study effect of additional confounding factors that affect neurological disability, such as cardiovascular risk factors. Incorporating such covariates may further strengthen models, but this will require detailed measurement of comorbidities and multi-organ functions and likely larger cohorts. 4. Some of the outcomes we used for modeling have not been extensively validated and are not used outside of our group, making it hard for readers to interpret. We already explained why such novel outcomes were necessary for successful modeling. Provided correlation matrices and recalibration of CSF biomarker-based models to predict traditional, well-known outcomes in the independent validation cohort successfully mitigated this limitation. 5. Correlation is not causation. Only successful interventional clinical trials can validate causality of disease mechanisms; however, such trials need a solid rationale. We believe this study provides such rationale. By applying creative analysis to differentiate likely candidate mechanisms from epiphenomena our study help prioritizing targets in future drug development.

Current study showcases that CSF biomarkers can quantify diverse molecular processes in living humans from very limited CSF sample and repeatedly. Such longitudinal measurements of cardiovascular risk factors were instrumental in determining their likely causality, linking their dose/exposure interactions to morbidity/mortality outcomes (43). Even though the proof of causality came from successful clinical trials (as it always must), measurements of biomarkers like LDL cholesterol selected trial population, measured pharmacodynamic effects and guided dose selections. Collectively, these studies led to understanding that multiple processes cause cardiovascular diseases, that they are not uniformly distributed among subjects, and if present, they must all be therapeutically inhibited by patient-specific, biomarker-guided polypharmacy regimen to effectively limit cardiovascular morbidity/mortality. We see strong parallels with our results: analogously to transformative pathology study revealing heterogeneity in the composition of acute MS lesions (44), we observed intra-individual heterogeneity in the non-lesional candidate pathogenic mechanisms. The patient-specific candidate causal mechanisms cannot be predicted by clinical classification of MS subtypes, they require CSF biomarker measurements. Because other than CNS-related pathways, like synaptogenesis, all remaining candidate causal mechanisms we identified are not organ specific, only CSF biomarkers can unequivocally link them to CNS tissue. While the assay we used is not clinical, we hope this study will spur interest in developing clinical-grade assays to comprehensively measure CSF biomarkers. Clinical-grade measurements of CSF biomarkers might greatly facilitate drug development for CNS diseases and eventually guide patient-specific polypharmacy regimen we take for granted in contemporary management of cardiovascular diseases, into neurological practice.

## MATERIALS AND METHODS

### Study design

This was a retrospective analysis of prospectively acquired cohort with details described in Figure 1, Table 1, and Supplemental Table 1.

### Subjects

A total of 394 MS patients (124 with PPMS, 179 with RRMS, and 91 with SPMS) and 45 HV were prospectively enrolled between January 1999 and December 2018 into natural history protocol “Comprehensive Multimodal Analysis of Neuroimmunological Diseases of the Central Nervous System” (Clinicaltrials.gov identifier NCT00794352); samples collected before 2009 were part of the “NIB Repository Protocol” (10-N-021). The protocol recruited patients with known or suspected diagnosis of MS. A thorough diagnostic workup included full neurological exam, MRI of the brain, functional tests (e.g., T25FW, nine-hole peg test [9HPT], SDMT, PASAT) and laboratory tests of blood and CSF. Each patient was followed for minimum of 1 year (with optional follow-up LP). Seventy-eight percent of CSF samples were collected in untreated stage. The inclusion criteria for HV cohort were between ages 18–75, lack of neurological diagnosis or systemic disease that would influence neurological functions or brain MRI and with vital signs in the normal range during the initial screening. The demographic data of all subjects are detailed in Table 1, demographic data for MS subtypes are in Supplemental Table 1.

### Sample processing

CSF was collected on ice and processed according to a written standard operating procedure by investigators blinded to diagnoses, clinical, and imaging outcomes. Aliquots were assigned alphanumeric identifiers and centrifuged for 10 minutes at 300g at 4°C within 30 minutes of collection. The pelleted CSF cells were processed by flow cytometry and the cell-free supernatants were aliquoted and stored in polypropylene tubes at −80°C.

### MRI imaging

MRI of the brain was generated on 1.5T and 3T scanners (General Electric & Siemens). T1 magnetization-prepared rapid gradient echo (MPRAGE) and T2-weighted 3D fluid attenuation inversion recovery (3D FLAIR) sequences were obtained. Assessment of contrast enhancement was performed using postcontrast (gadopentetate dimeglumine at 0.1 mmol/kg) T1-weighted and postcontrast FLAIR images. The number of CELs was recorded in the research database. The MRI protocol extended caudally to C5 level to allow analysis of upper cervical SC.

The volumetric analysis was performed using LesionTOADS volume segmentation algorithm performed on QMENTA imaging platform (www.qmenta.com). The details of the analyses have been described (45). BPFr was calculated as a ratio between the total brain volume and intracranial cavity volume. Upper cervical SC cross-sectional area (C1–C2) was calculated from brain MRI images using Spinal Cord Toolbox (46). The investigators generating volumetric MRI data were blinded to diagnostic codes, clinical outcomes or any laboratory outcomes.

### Clinical outcomes

Neurological exams have been recorded directly (after 9/2017) or transcribed retrospectively from structured medical records neurological examination form (before 9/2017) into NeurEx™ App (47) that automatically calculates traditional MS disability outcomes –EDSS (9), SNRS (19), Hauser AI (21), IPEC disability scale (20). CombiWISE was calculated from EDSS, SNRS, T25FW and nondominant hand of 9HPT, as described (14). EDSS-based MS severity outcomes –MSSS (48), ARMSS (10), and MS-DSS (17) - were calculated as described.

Generation of novel outcomes for this study are described in the Results section.

### Flow cytometry

Fresh CSF cells collected from ~20cc of CSF were resuspended in 200ul of X-VIVO™ 15 (Lonza, REF: 04-418Q) on ice. A minimum of 2,000 cells were stained with a 12-color antibody panel, analyzed by BD Bioscience LSR II flow cytometer, and gated using BD FACSDiva software as described (49). The results have been prospectively entered into the research database, quality-controlled and locked from further changes. The database automatically calculated proportions (percentages) of different cell populations based on total cell number.

### CSF immunoassays

CSF levels of NFL, CD27, CHI3L1, and SERPINA3 were measured by personnel blinded to diagnostic, clinical and MRI outcomes. NFL and SERPINA3 were quantified using spectrophotometric assays by UmanDiagnostics (catalog# 10-7002) and RayBiotech (catalog# ELH-SERPINA3-1) respectively. CHI3L1 and CD27 were quantified via homebrew assays on MSD-ECL platform using antibodies from R&D systems (catalog# DY2599) and Sanquin (catalog# M1960) respectively. Details of the assays have been published previously (50).

### SOMAScan analysis

Total of 1,042 unique CSF samples marked with alphanumeric coded were analyzed by SOMAScan (Somalogic Inc, Boulder, CO, USA). Samples representing different diagnostic groups were interspersed between individual 96-well plates, while longitudinal samples of a single patient were kept withing the same plate. SOMAScan assay measured relative fluorescent units (RFU) of 5,034 DNA aptamers, called SOMAmers. The raw RFUs have been mathematically processed to normalize the hybridization signal within each plate, and to calibrate the signal across different plates, using control samples embedded within each plate.

### Adjustment for physiological aging and sexual dimorphism

To regress out the effect of natural aging and sexual dimorphism for each SOMAmer the normalized and calibrated RFUs were log-transformed and a linear regression model using age and sex as two independent variables was generated in the HV cohort. The prediction of this model was then subtracted from each sample, resulting in age- and sex-adjusted RFUs, with mean of HV cohort of zero. For further analyses, the SOMAmer levels of MS patients were also standardized to HV, resulting in HV mean of zero and HV SD of one.

### Ingenuity Pathway Analysis (IPA^®^)

To identify biological processes associated with MS outcomes, first, we performed a univariate correlation between outcomes (T2LL, BD, CombiWISE, SC disability for MS progression and BD severity, CombiWISE severity, SC severity, MS-DSS for MS severity) and age-/sex-adjusted SOMAmers in the MS cohort. For statistically significant correlations (FDR-adjusted p < 0.05) the Spearman Rho coefficients were submitted into IPA^®^ and a Core Expression Analysis was performed using “Expr Other” as a measurement type. The identified Canonical pathways, Upstream regulators, and Causal networks were exported for each outcome and merged using RStudio software Version 1.1.463 (R version 4.0.2) (51). A sum of z-scores and sum of −log_10_ *p*-value were calculated for each pathway/upstream regulators/causal network across the four MS progression and four MS severity outcomes.

For longitudinal changes, pairs of untreated CSF samples were identified maximizing the length of follow-up. The differences in age-/sex-adjusted SOMAmer levels between last and first LP were calculated for each patient. Paired one-sample two-sided Wilcoxon test identified statistically significant changes (rejecting the null hypothesis of no change over time) after the FDR adjustment for multiple comparisons. The median values of significant longitudinal changes were then submitted to IPA^®^ followed by the same analysis as described above.

MS-specific differences were calculated between HV cohort and MS cohort represented by first LP per patient. The median levels of SOMAmers in the HV cohort were subtracted from median levels of MS patients and unpaired two-sample two-sided Wilcoxon test was used to identify statistically significant differences between MS and HV that were again analyzed by IPA^®^ as described above.

To identify biology that differentiates brain vs SC damage, we generated two cohorts of propensity score matched samples: 1) Linear regression model between BD and CombiWISE identified CombiWISE residuals—patients with proportionally higher and lower measured CombiWISE than what would be predicted by their BD levels. Patients with CombiWISE residuals within the interquartile range (IQR) of the data distribution were removed and remaining patients with residuals less than Q1 and greater than Q3 were matched for BD levels using propensity score matching (matchit function with “full” method; “MatchIt” R package (52)). Differences in age-/sex-adjusted SOMAmers between matched samples were assessed by paired two-sample two-sided Wilcoxon test after FDR-adjustment for multiple comparison. The medians of differences for identified SOMAmers were analyzed by IPA^®^. 2) The same workflow was applied to comparison between BD and CombiWISE, by regressing out disability measured by CombiWISE and calculating BD residuals.

To identify differences in biology representing MS progression vs severity, the canonical pathways *z*-scores and −log_10_ *p*-values were summed up across the four MS progression and four MS severity outcomes. Differentially activated pathways were identified as those with absolute sum of *z*-scores greater than 4, sum of −log_10_ *p*-value greater than 1.33 and the difference between sum of *z*-scores for severity and sum of *z*-scores for progression greater than 2.

To calculate patient-specific pathways activation scores, the details for each canonical pathway were downloaded from IPA^®^ and intersected with proteins measured by SOMAscan. The HV-normalized SOMAmer values corresponding to pathway components and measured at first untreated CSF sample were averaged, after adjustment for their predicted effect on pathway activation (e.g., adding scores of analytes predicted to activate the pathway and subtracting scores of analytes predicted to inhibit the pathway). The patient-specific pathway scores were analyzed by unsupervised hierarchal clustering using the “ward.D” clustering as part of the “Pheatmap” R package (https://CRAN.R-project.org/package=pheatmap).

### Machine learning models

To formally test whether our training cohort was sufficiently powered to find optimal solution with numbers of predictors exceeding numbers of subjects, two sets of predictors were considered, 5,034 age- and sex-adjusted single SOMAmers, and 12.9 million age- and sex-adjusted SOMAmer ratios. Two modeling strategies were tested: EN (linear modeling strategy that can handle collinearity between some SOMAmers) and RF (a tree-based algorithm capable of capturing nonlinear relationships and disease heterogeneity). Models were generated in randomly split two-thirds of MS patients that constituted the “training cohort” and the performance of the models was tested in the remaining one-third of the MS patients that constituted the “validation cohort”. The models were generated either using all available LPs per patient (i.e., including CSF from treated patients) in the training cohort or using only the first LP per patient. The validation was performed in both cases using only the first LP per patient in the validation cohort.

The EN models on single SOMAmers were generated using glmnet R package (53) and the EN models on SOMAmer ratios utilized biglasso R package (54); the RF modeling used ranger R package (55). For RF models an iterative process of dimension reduction was performed, as described (56), by removing bottom 10% of variables with the least variable importance measured based on node impurity. The process was repeated until root mean square error (RMSE) stabilized or increased. The OOB error was assessed for each final RF model. Spearman correlation coefficient (Rho), coefficient of determination (R^2^), CCC, and RMSE were calculated to assess the models’ performance.

### Statistics

All data were analyzed using R Studio software (details of packages used for analyses are mentioned above). Statistical analyses were performed using one- or two-sample two-sided Wilcoxon tests or t-test. FDR-adjustment was used to correct for multiple comparisons. More details on statistical methods used are described above.

### Study approval

Clini

## Data Availability

The datasets generated during and/or analyzed during the current study as well as the custom codes used to perform analyses will be available from the corresponding author after the manuscript has been peer-reviewed and published.

## Abbreviations

9HPT: nine-hole peg test
AI: ambulation index
ARMSS: age-related multiple sclerosis severity
BBB: blood-brain barrier
BD: brain damage
BPFr: brain parenchymal fraction
CCC: concordance correlation coefficient
CEL: contrast-enhancing lesion
CombiWISE: combinatorial weight-adjusted disability score
CNS: central nervous system
CSF: cerebrospinal fluid
DC: dendritic cell
EAE: experimental autoimmune encephalomyelitis
EBV: Epstein-Barr virus
EDSS: expanded disability status scale
EMT: epithelial-mesenchymal transition
EN: elastic net
FDR: false discovery rate
FLAIR: fluid attenuation inversion recovery
HV: healthy volunteer
IPA: Ingenuity Pathway Analysis
IPEC: Instituto de Pesquisa Clinica Evandro Chagas
IQR: interquartile range
lncRNA: long non-coding RNA
LP: lumbar puncture
MPRAGE: magnetization-prepared rapid gradient echo
MRI: magnetic resonance imaging
MS: multiple sclerosis
MS-DSS: multiple sclerosis disease severity scale
MSSS: multiple sclerosis severity score
NIH: National Institutes of Health
NK cell: natural killer cell
NO: nitric oxide
OOB: out-of-bag
PASAT: paced auditory serial addition test
PIRA: progression independent of relapse activity
PPMS: primary progressive multiple sclerosis
PMS: progressive multiple sclerosis
PRR: patter-recognition receptors
RF: random forest
RFU: relative fluorescent units
RMSE: root mean square error
ROS: reactive oxygen species
RRMS: relapsing-remitting multiple sclerosis
SC: spinal cord
SDMT: symbol-digit modalities test
SNRS: Scripps neurological rating scale
SPMS: secondary progressive multiple sclerosis
T25FW: timed 25-foot walk
T2LL: T2 lesion load

## List of Supplementary Materials

Supplementary results

Supplemental Figure 1

Supplemental Figure 2

Supplemental Figure 3

Supplemental Figure 4

Supplemental Figure 5

Supplemental Figure 6

Supplemental Table 1

Supplemental Table 2

Supplemental Table 3

Supplemental Table 4

Supplemental Table 5

## Acknowledgments

This study was conducted as part of Cooperative Research and Development Agreement between National Institutes of Health, Novartis, and Somalogic.

The content of this publication does not necessarily reflect the views or policies of the Department of Health and Human Services, nor does mention of trade names, commercial products, or organizations imply endorsement by the U.S. Government.

The authors would like to thank Yolanda L. Jones, NIH Library, for editing assistance.

This work utilized the computational resources of the NIH HPC Biowulf cluster (http://hpc.nih.gov).

## Author contributions

Conceptualization: BB

Methodology: PK, KL, JW, CJL, BB

Software: PK, KL, JW, CJL

Formal analysis: PK, KL, JW

Investigation: PK, RM, YK, MV, BB

Visualization: PK, BB

Writing – original draft: PK, BB

Writing – review & editing: PK, KL, JW, CJL, RM, YK, MV, LJ, BB

Funding acquisition: BB, LJ

