## Supplemental material for "Molecular mechanisms associated with multiple sclerosis progression, severity and phenotype"

### Supplemental materials

#### Supplemental results – CSF biomarker-based models of MS progression and severity

To investigate if modeling strategy that captures non-linear relationships between the CSF biomarkers outperforms linear models, we compared elastic net (EN) with random forest (RF) algorithms. We expected that RF models based on SOMAmer ratios outperform other modeling strategies. Although there was no dramatic difference in performance of EN and RF models, we concluded that CSF biomarkers contribute to outcome prediction linearly, with low effect sizes, requiring integration of hundreds, even thousands of measurements. In this regard the RF models are generally more efficient: achieving comparable effect sizes with much smaller number of variables.

Next, we tested the hypothesis whether increased number of samples in the training cohort improves model performance in the validation cohort. We achieved this by using the first available sample for each training cohort patient only, and by including all available samples for training cohort patients (including longitudinal samples). The performance of these models was tested in the same validation cohort comprised of one sample per patient ($1^{st}$ available LP). As shown in Fig. S4, having more training data did enhance validation performance, but only marginally. This indicates that existing size of our modeling cohort was sufficient to reliably capture biology measured by 5k SOMAScan that underlie development of these nine modeling outcomes. Our results predict that expanding the number of CSF biomarkers measured (provided they capture non-overlapping and relevant biology), rather than expanding cohort sizes, will lead to stronger models.

The last modeling aspect we tested was pre-filtering. We have shown previously that models built from SOMAmer ratios outperform models built from single individual SOMAmers (42). However, increased number of predictors means higher computational power demand. To keep the computational time required to build the models at a reasonable level, in the past we used signal-to-noise analysis to restrict SOMAmer ratios input, although it remained unclear whether such pre-filtering step decreased models' performance. Consequently, here we used NIH HPC Biowulf cluster (www.hpc.nih.gov) to generate models from 12.9M variables. These models were so computationally demanding that their generation and optimization took weeks for each outcome.
To test the hypothesis whether noise filtering in predictors effects model performance in the validation cohort we built the EN models using the full set of 5,034 SOMAmers and in parallel, built the models using a reduced set of predictors. The filtering was achieved by eliminating SOMAmers with interclass correlation coefficient (ICC) lower than 0.5, effectively eliminating 49% of SOMAmers. The ICC analysis uses linear mixed-effect model to calculate how much variance is explained by random effect – in our case, identifying SOMAmers that are relatively stable over time (longitudinal samples) and vary among patients (cross-sectional samples). We have observed that the models that employed pre-filtering of biomarkers based on signal-to-noise ratio had comparable independent validation performance with models that used all available biomarkers (Fig. S5). Considering how computationally demanding these models were, prefiltering step is highly advisable.

Lastly, we compared all nine models (using all different modelling strategies) and found out that: 1) different algorithms results in similar set of SOMAmers (or SOMAmers in ratios) selected by each model for a particular outcome, 2) A relatively small subset of SOMAmers is represented across all outcomes/models, most likely representing the high-signaling SOMAmers that differentiates patients with different levels of MS progression/severity. For details, see Table S5.

#### Supplementary Figures

#### Supplemental Figure 1

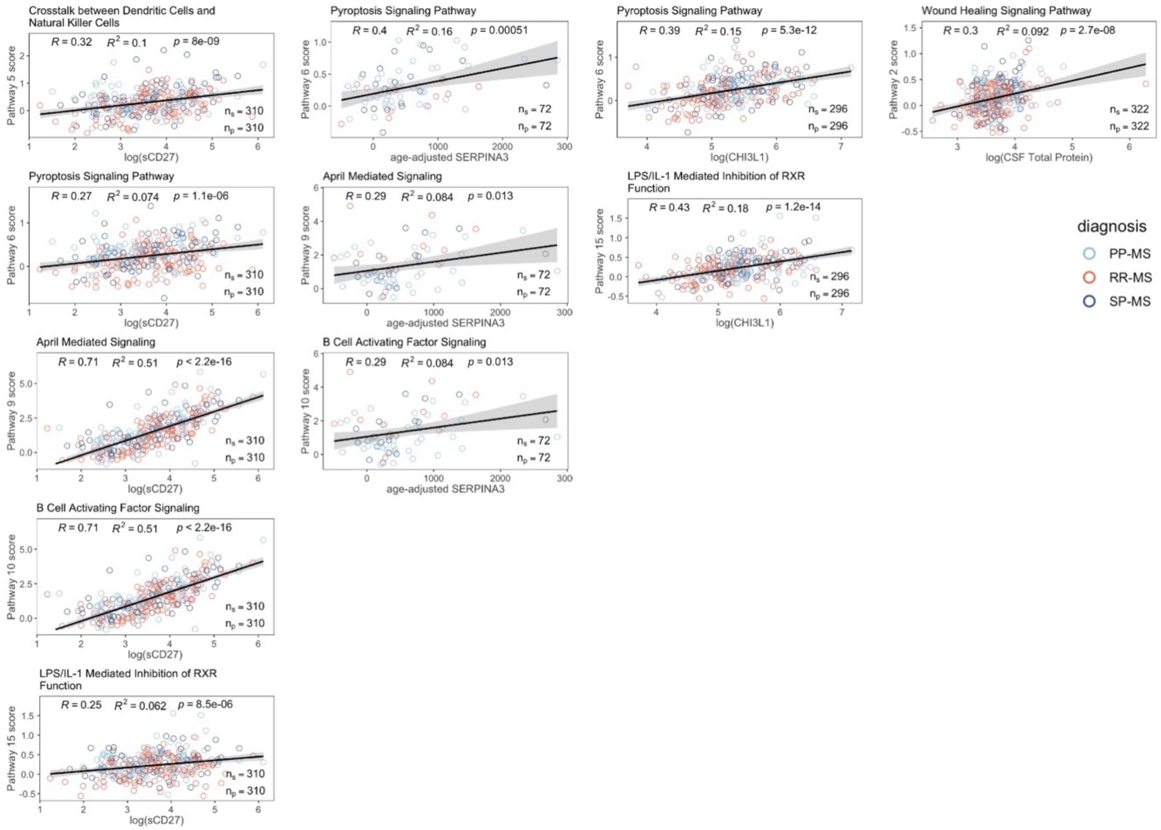


**Supplemental Figure1. Correlation of selected CSF biomarkers with pathway scores associated with MS severity.** A set of biomarkers (sCD27, CHI3L1, SERPINA3, and total protein) was measured in the CSF of untreated MS patients. Correlations between these biomarkers and pathways scores associated with MS severity are displayed. The correlation was evaluated by Pearson R, coefficient of variance ($R^2$) and p-value. No adjustment for multiple comparison was performed.

**Supplemental Figure 2**

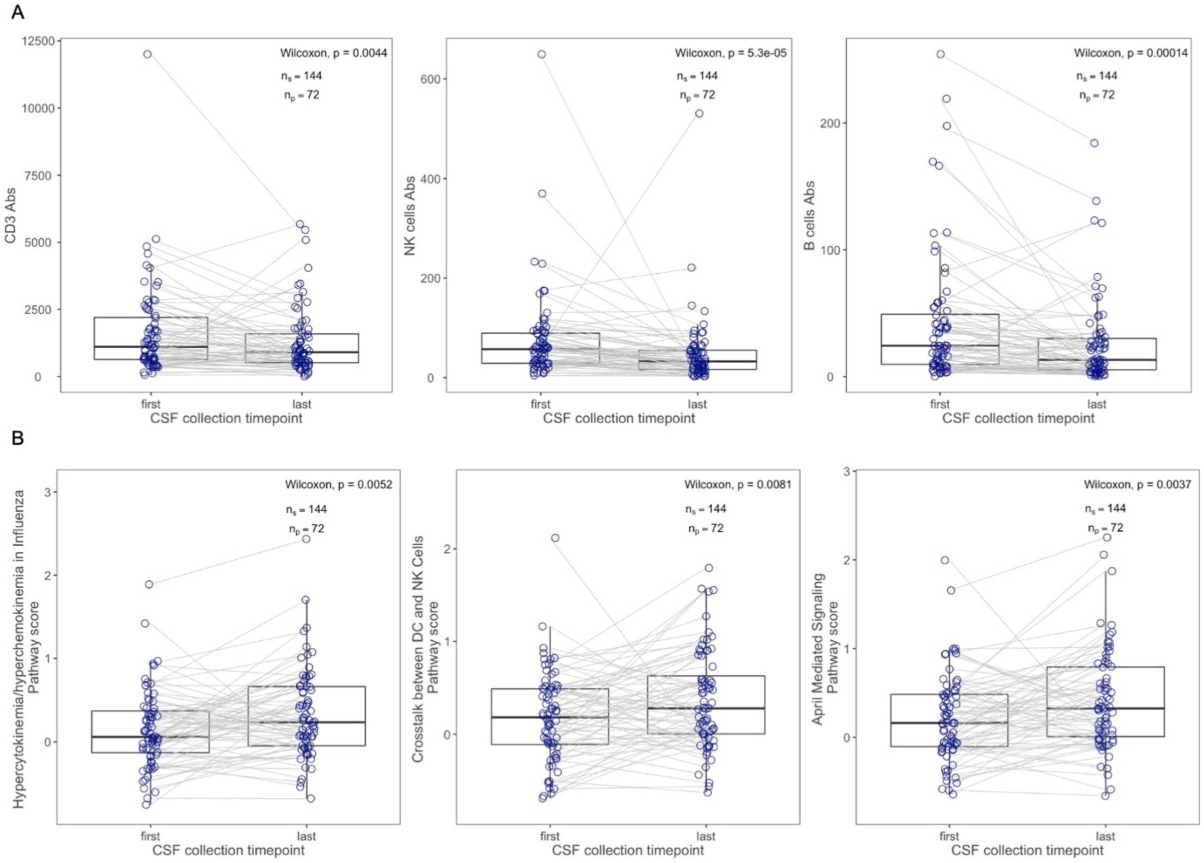


**Supplemental Figure 2. Comparison of absolute CSF cell counts and progression pathway scores in the untreated longitudinal cohort of MS patients.** (A) Immune cells isolated from longitudinal CSF samples collected over at least 1 year follow-up in untreated MS patients were analyzed by flow cytometry – examples of quantified lymphocytes (CD3+), NK cells, and B cells all show significant decrease over time. (B) In contrast to absolute cell numbers, scores for pathways associated with MS progression show statistically significant increase over time in the same cohort of untreated MS patients.

### Supplemental Figure 3

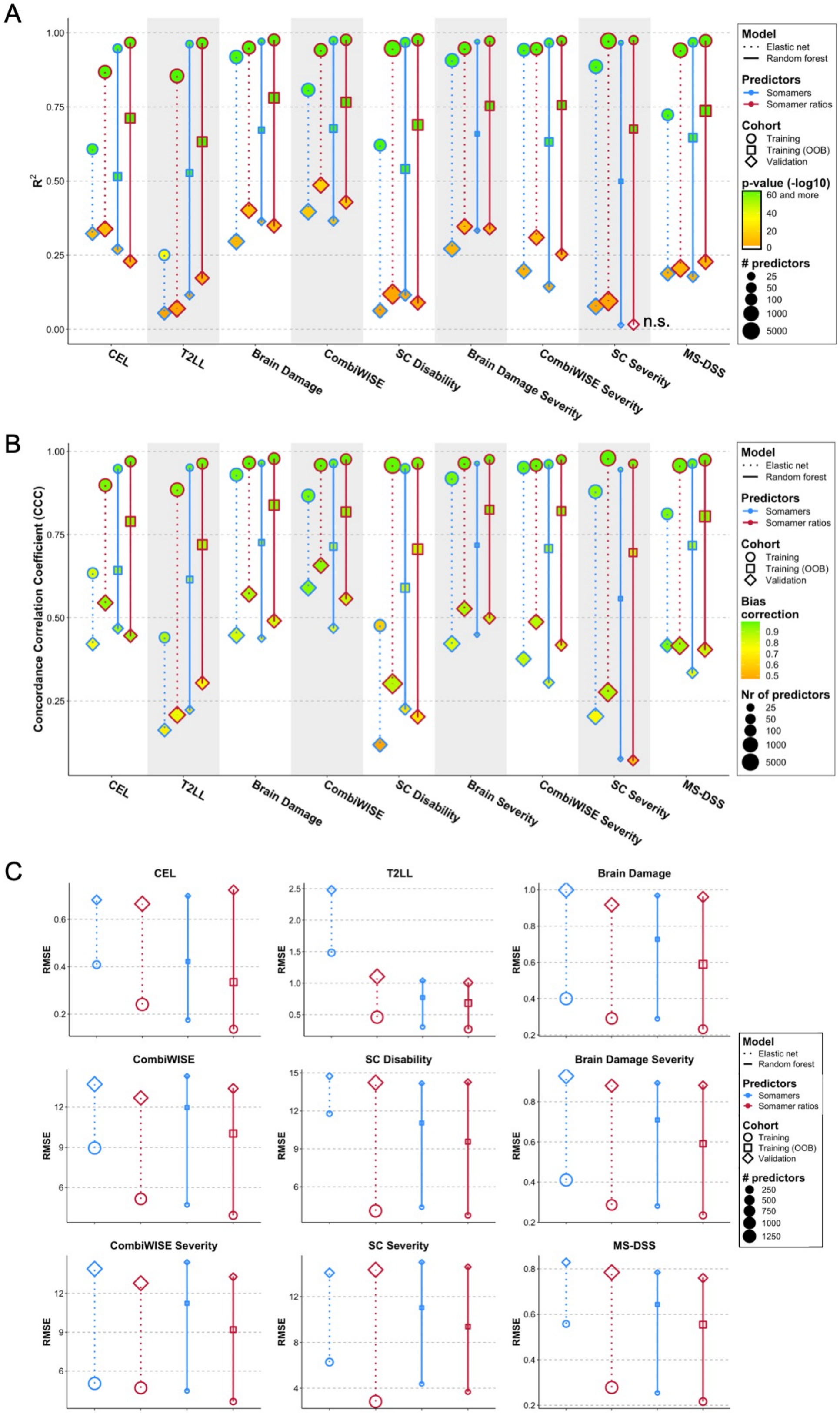

**Supplemental Figure 3. Characteristics of CSF-based models of MS outcomes.** (A) A comparison of models' performance based on Coefficient of variance ($R^2$) in training (circles), OOB (squares), and validation (diamonds) cohorts. The EN models are illustrated by dotted lines connecting training and validation cohort sets, while RF models are shown connected with solid lines. Models built with single SOMAmers as predictors are in blue, models generated from SOMAmer ratios are shown in red. The size of the points represents the number of predictors retained in the final model, the fill of the datapoints illustrates $-\log_{10}$(p-value) of the linear regression model between predicted and measured outcomes. Two RF models of SC severity did not validate (n.s.). (B) A comparison of models' performance based on Concordance Correlation Coefficient (CCC). The fill of the points represents the Bias correction – a coefficient determines the deviation of the linear regression slope from the 1:1 line (slope of 1). (C) Root-mean-square error (RMSE) – a standard deviation of the residuals (prediction errors) among four different modelling strategies for 9 MS outcomes.

**Supplemental Figure 4**

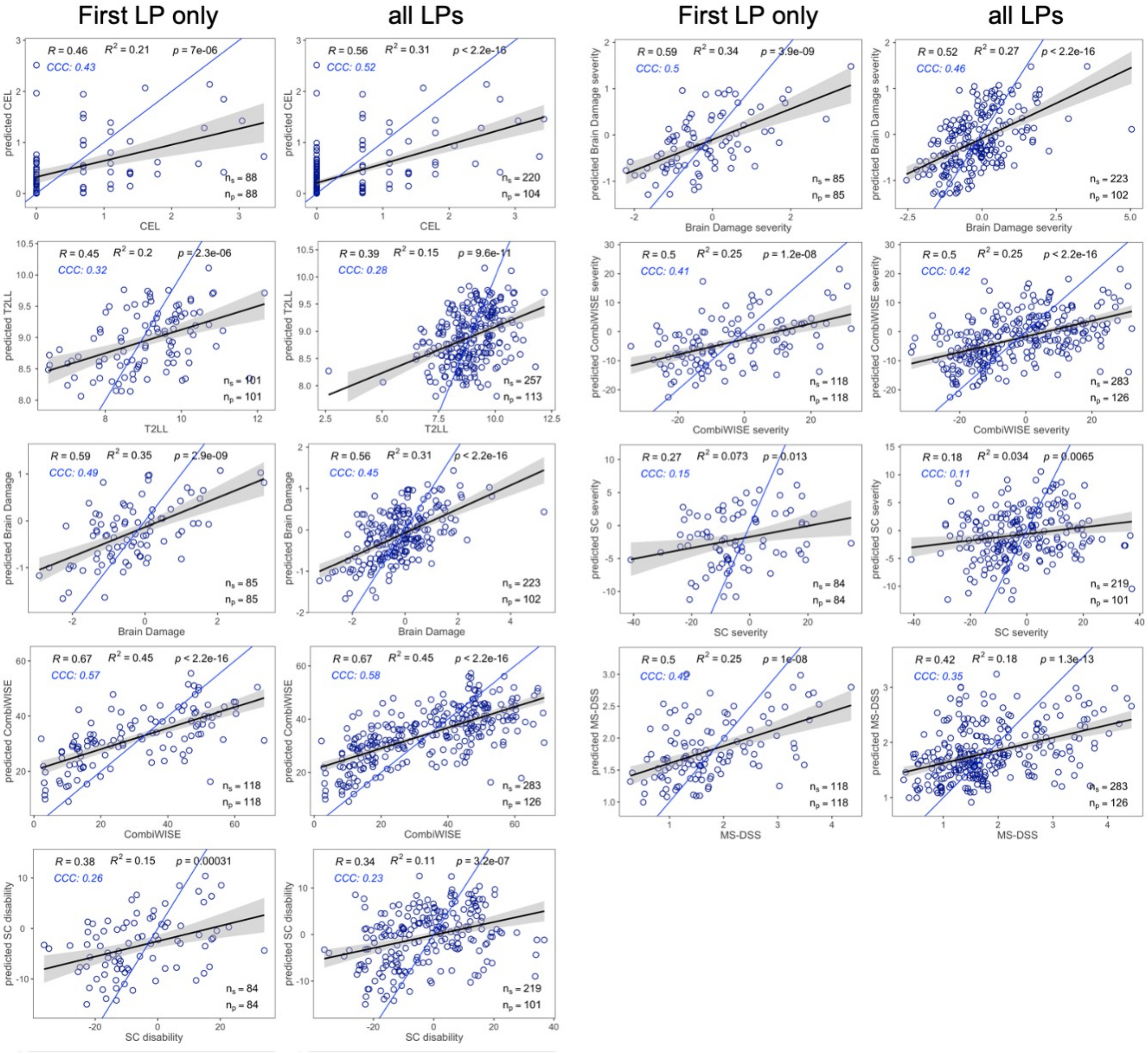


**Supplemental Figure 4. Validation results – first sample vs all samples.** The performance of RF models built from SOMAmer ratios in the training cohort of samples were tested in validation cohort comprised of "First LP only" – first available longitudinal sample per patient – and of "all LPs" – all available longitudinal samples for validation cohort patients. The performance of the models was evaluated by Pearson R, coefficient of variance ($R^2$), p-value, and concordance correlation coefficient (CCC). X-axis shows measured outcomes, y-axis shows predicted outcomes. The black line represents a fitted line of a simple linear regression model with 95% confidence interval represented by the gray area. Blue line represents 1:1 line – a perfect fit between measured and predicted values. $n_s$ – number of samples, $n_p$ – number of patients.

**Supplemental Figure 5**

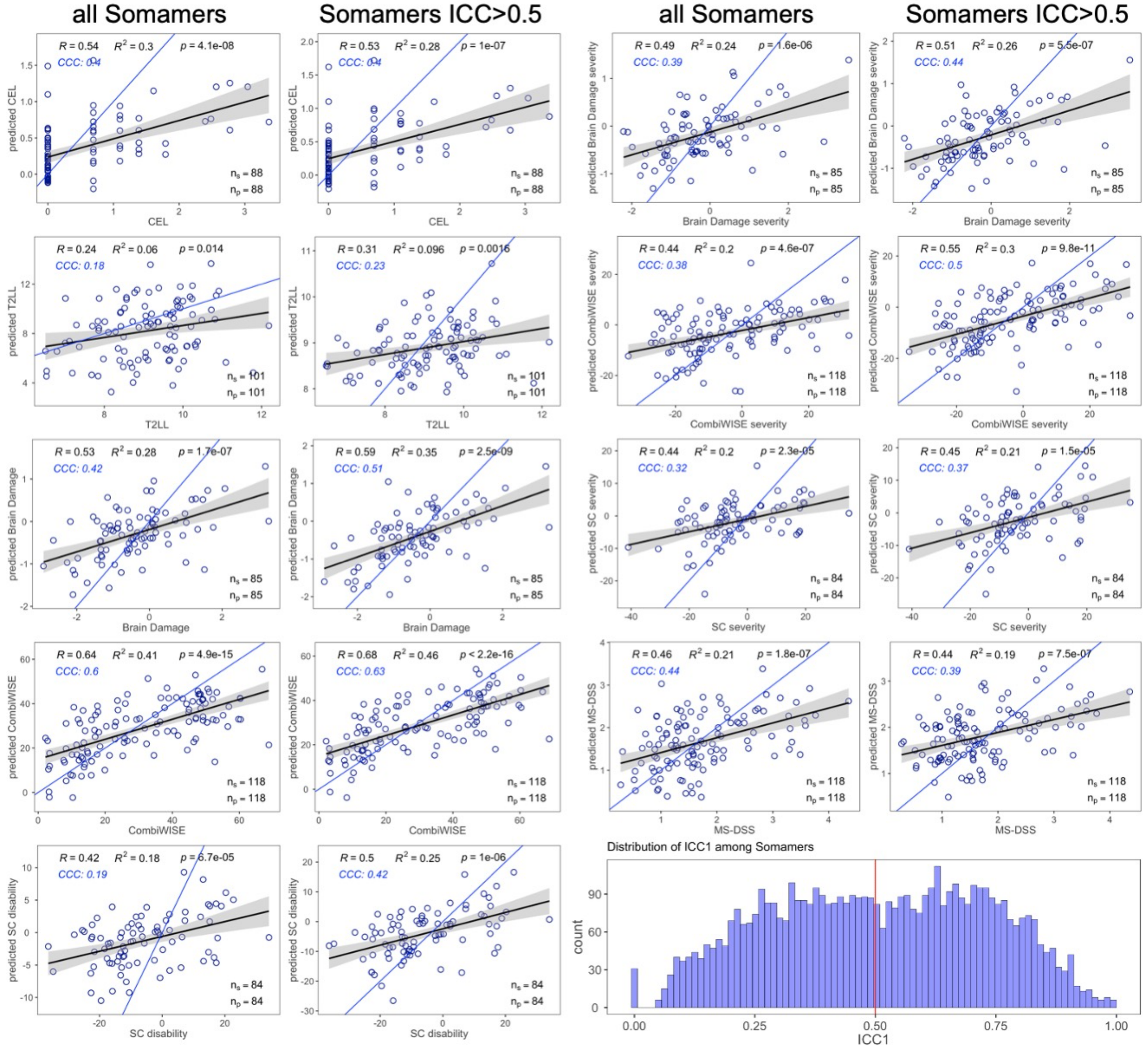


**Supplemental Figure 5. Validation results – full vs reduced predictor dataset.** The performance of the EN models built from individual SOMAmers in the training cohort was tested by using "all SOMAmers" – all measured 5034 SOMAmers – and by using "SOMAmers ICC>0.5" – a subset of SOMAmers with high signal-to-noise ratio. The Interclass Correlation Coefficient (ICC) for each SOMAmer was calculated in the dataset of all MS and HD samples by generating a two-way mixed-effect models with patient ID as a random effect. Only SOMAmers with ICC > 0.5 were used to build the models. The performance of the models was evaluated by Pearson R, coefficient of variance ($R^2$), p-value, and concordance correlation coefficient (CCC). X-axis shows measured outcomes, y-axis shows predicted outcomes. The black line represents a fitted line of a simple linear regression model with 95% confidence interval represented by the gray area. Blue line represents 1:1 line – a perfect fit between measured and predicted values. $n_s$ – number of samples, $n_p$ – number of patients. The histogram in the lower right corner shows distribution of ICCs across all 5,034 SOMAmers, the red vertical line marks the ICC>0.5 cut-off.

**Supplemental Figure 6**

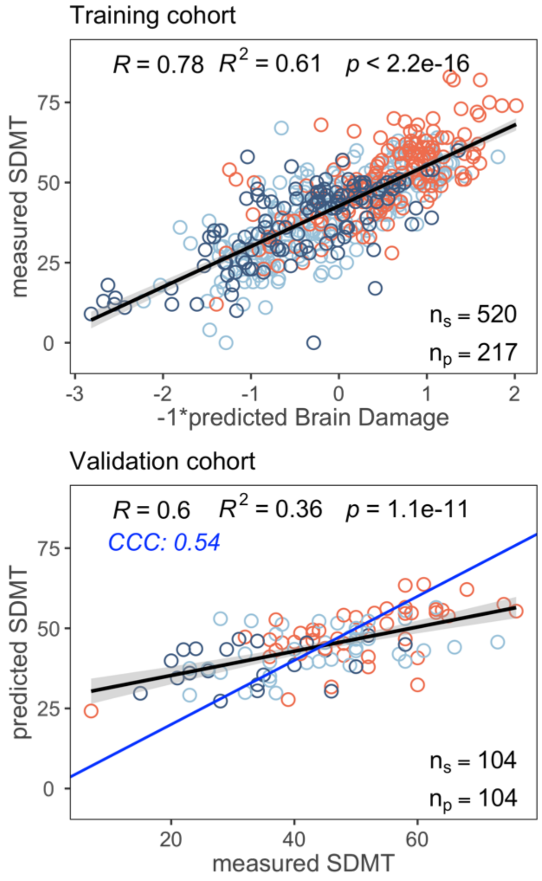


**Supplemental Figure 6.** SOMAmer-predicted disability outcome Brain Damage was used to generate a simple linear regression model of measured SDMT in the training cohort (top plot). The regression equation was then used in the independent validation cohort to predict SDMT, reaching CCC of 0.54 (bottom plot). The blue line represents 1:1 line of perfect fit between measured and predicted outcome.

#### Supplemental Tables

**Supplemental Table 1. Demographic data for MS subtypes**

**Supplemental Table 2. Correlation between MS disability/severity outcomes and age/sex-adjusted SOMAmers**

**Supplemental Table 3. Upstream regulators and Causal networks enriched in severity outcomes compared to progression outcomes**

**Supplemental Table 4. Models' performance in training and validation cohorts**

**Supplemental Table 5. Variable importance across different ML models**
